# Recombinant VEGF-C (Cys156Ser) protein restores mesenteric lymphatic drainage and improves gut immune surveillance in experimental liver cirrhosis

**DOI:** 10.1101/2020.12.24.20248815

**Authors:** Pinky Juneja, Syed Nazrin Ruhina Rahman, Dinesh M Tripathi, Impreet Kaur, Sumati Rohilla, Abhishek Gupta, Preety Rawal, Sukriti Baweja, Archana Rastogi, VGM Naidu, Shiv K Sarin, Subham Banerjee, Savneet Kaur

## Abstract

**Background and Aim:** Lymphatic vessels (LVs) are crucial for maintaining abdominal fluid homeostasis and immunity. In liver cirrhosis, mesenteric LVs (mLVs) are dilated and dysfunctional. Given an established protective role of VEGF-C in LVs, we hypothesized that VEGF-C treatment could improve functions of mLVs in cirrhosis.

**Method:** In this study, we developed a nanoformulation comprising LV-specific growth-factor, recombinant human VEGF-C(Cys156Ser) protein(E-VEGF-C) and delivered it orally in rat models of liver cirrhosis to target mLVs. Nanoformulation without VEGF-C served as vehicle. Drainage of mLVs was analyzed using tracer dye. Portal and systemic physiological assessments and computed-tomography were performed to measure portal pressures and ascites. Gene expression of mesenteric lymphatic endothelial cells (LyECs) was studied. Immune cell subsets in mesenteric lymph nodes (MLNs) were quantified by flow-cytometry. Gut bacterial translocation to MLNs was examined using GFP-labelled bacteria.

**Results:** In cirrhotic rats, mLVs were dilated and leaky with impaired drainage. Treatment with E-VEGF-C induced proliferation of VEGFR3+ mLVs, reduced their diameter and improved functional drainage. Ascites and portal pressures were significantly reduced in E-VEGF-C treated rats compared to vehicle. At molecular level, E-VEGF-C treatment upregulated the expression of cell adhesion and permeability genes (VCAM1, VE-Cad) in LyECs. In MLNs of E-VEGF-C rats, there was an increased percentage of CD8+CD134+ T-cells and decreased CD25+Treg-cells. Bacterial translocation was also limited to MLNs only in E-VEGF-C treated rats with reduced levels of endotoxins in ascites in comparison to vehicle.

**Conclusion:** E-VEGF-C treatment ameliorates mesenteric lymph drainage, portal pressure, and strengthens cytotoxic immune responses in MLNs in experimental cirrhosis. It may thus serve as a promising therapy to manage ascites and portal pressure and reduce gut bacterial translocation in patients with cirrhosis.

**Lay Summary:** A human recombinant pro-lymphangiogenic growth factor, VEGF-C, was encapsulated in nanolipocarriers (E-VEGF-C) and orally delivered in rat models of decompensated liver cirrhosis to facilitate its gut lymphatic vessel uptake. E-VEGF-C administration significantly increased mesenteric lymphatic vessel proliferation and improved lymph drainage, attenuating abdominal ascites and portal pressures in the animal models. E-VEGF-C treatment limits bacterial translocation to MLNs only with reduced gut bacterial load and ascitic endotoxins. E-VEGF-C therapy holds the potential to manage ascites and portal pressure and reduce gut bacterial translocation in patients with decompensated cirrhosis.

**Graphical Abstract:** 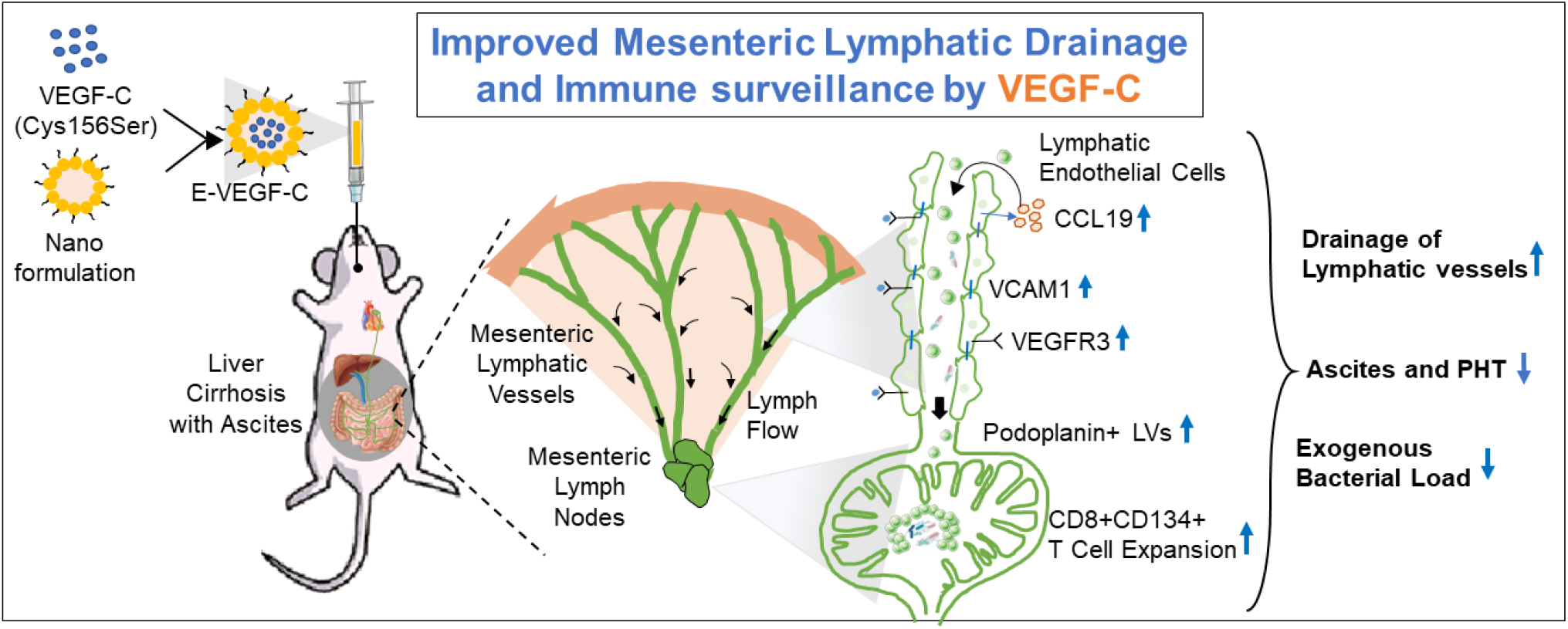

## Introduction

Chronic liver disease is associated with significant morbidity and mortality worldwide[1,2]. They can progress from mild fibrosis to cirrhosis, which may further progress to end-stage liver disease with the onset of decompensation such as ascites, hepatic encephalopathy, and variceal bleeding[3]. In addition to other underlying factors, deranged gut-liver axis plays significant role in the progression of liver disease[4,5]. Whatever comes from the gut enters liver through portal circulation before returning to the heart and systemic blood via hepatic vein. However, lymphatic channels in the gut bypass the portal circulation in liver and directly connect the gut to systemic circulation via thoracic duct[6].

Gut lymphatic vessels (LVs) play essential roles in the absorption and transportation of fat, maintenance of fluid homeostasis, removal of interstitial fluid, macromolecules, immune cells, and microbial debris from intestine, and returning lymph to the blood circulation, thereby preventing edema and abdominal ascites[7]. Lymphangiogenesis (proliferation of LVs from the pre-existing LVs) in the gut during inflammation compensates for lymphatic insufficiency in patients with inflammatory bowel disease (IBD), such as Crohn’s disease, where there is both an increase and a redistribution of LVs[8,9]. In liver cirrhosis, the production of abdominal lymph increases 30-fold, and there is a positive correlation between lymph flow and increasing portal pressure[10,11]. Using animal models of CCl4-induced liver damage, an elegant study reported that gut LVs show an impaired phenotype and reduced contractility in cirrhosis[12].

Vascular endothelial growth factor-C (VEGF-C), key pro-lymphangiogenic factor, activates lymphangiogenesis by binding to tyrosine kinase receptor VEGF receptor-3 (VEGFR3)[13]. Therapies aimed at enhancing lymphangiogenesis and improving lymphatic function and transport with VEGF-C are effective for improving lymphatic drainage and ameliorating inflammation in diseases such as IBD, rheumatoid arthritis, skin inflammation, and hepatic encephalopathy[14–17]. However, the short half-life and systemic side effects of VEGF-C are major hurdles that limit their clinical application[18].

Here, we hypothesized therapeutic role of VEGF-C for the restoration of gut lymphatic drainage and immune response in cirrhosis. VEGF-C, a hydrophilic molecule, carries positive charge at physiological pH with short half-life and is highly unstable in nature[19,20]. To minimize the side effects of systemic delivery and increase the gut bioavailability of VEGF-C, we fabricated an engineered VEGF-C protein using human recombinant VEGF-C protein (Cys156Ser), which specifically binds to VEGFR3 homodimers, present majorly on Lymphatic Endothelial cells (LyECs) lining the LVs and encapsulates it within nanoscale reverse micelle (RM)-based lipocarriers and delivered it via oral route to ensure lymphatic uptake for targeted gut lymphangiogenesis in vivo.

## Materials and Methods

### Preparation of E-VEGF-C

To prepare reverse micelles (RMs), high-pressure homogenization/micro-fluidization techniques were adapted as described in this [21], followed by slight modification. Recombinant human VEGF-C (Cys156Ser) was incorporated in prepared RMs to obtain nanosized RMs containing VEGF-C protein termed as engineered VEGF-C (E-VEGF-C) **(Supp-Figure 1)**. Further characterization of E-VEGF-C is provided in supplementary materials.

**Figure 1:**
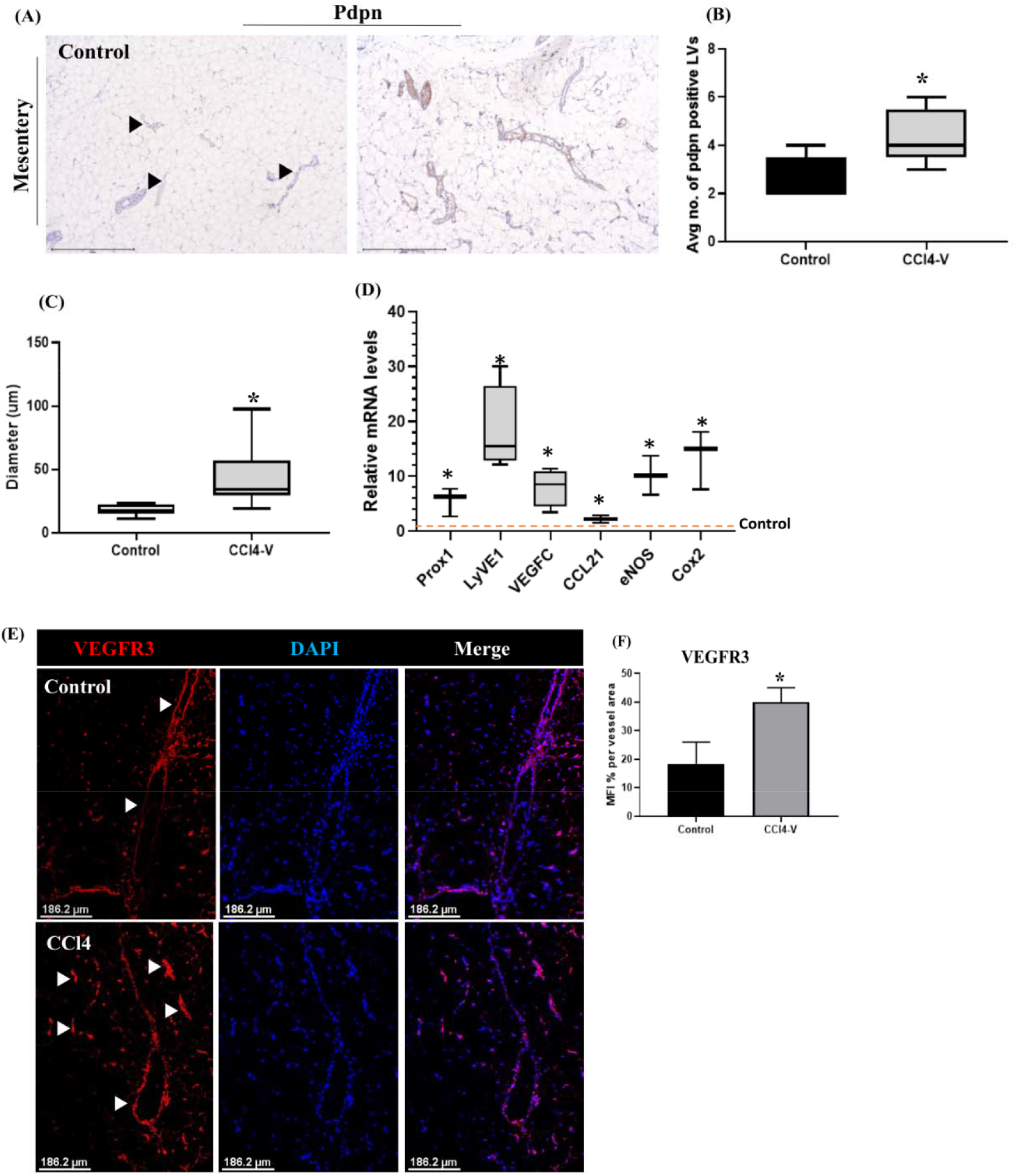
Mesenteric Lymphatic vessels (mLVs) density increased in liver cirrhosis. **(A)**Immunohistochemical staining with podoplanin (pdpn) antibody was performed in mesentery tissue of control and CCl4 animal model of liver cirrhosis. Scale bar:500μm. **(B)**The number of the pdpn+ mLVs was quantified using ImageJ. *P*=0.0240. **(C)**The diameter of mLVs in control and CCl4 rats was measured using ImageJ’s diameter plugin. n=5 each. *P*=0.0119 (**D)**mRNA level for Prox1, LyVE1, VEGF-C, CCL21, eNOS, and COX2 were quantified in mesentery tissue using qRT-PCR. n=5 each. Two-tailed unpaired T-test. *P*=0.05-0.01, dotted line represents control. **(E)**Immunofluorescence staining for VEGFR3 antibody and DAPI was performed in mesentery tissue of control and CCl4 rats. Scale bar:186.2μm. **(F)**VEGFR3 expression was quantified from the same sections using ImageJ. n=5 each. Data is expressed as mean±SD. Two-tailed, unpaired T-test, *P*=0.0147.’*’ represents p< 0.05.

### Study Groups and treatment

All animals received humane care according to the criteria outlined in the ‘‘Guide for the Care and Use of Laboratory Animals’’ by the National Academy of Sciences and published by the National Institutes of Health (NIH publication 86-23 revised 1985). The study was approved by the Animal Ethics Committee of ILBS, New Delhi, according to standard guidelines (Ethics Protocol No: IAEC/ILBS/18/01). Rats were housed in a room at 22 ± 3^0^C for a 12 hrs. light-dark cycle and were given food and water ad libitum. Studies were performed on 8-week-old male Sprague Dawley (SD) rats weighing 250–300g. Two separate models for portal hypertension were prepared: (1) cirrhotic models of portal hypertension (2) non-cirrhotic portal hypertension models. Cirrhosis was induced by intraperitoneal injection of 1.0 ml/kg body weight of CCl4:olive oil in 1:1 ratio, two times a week for 12-14 weeks until ascites formation. Non-cirrhotic animal models of portal hypertension were developed using partial portal vein ligation (PPVL), as previously described [22]. Briefly, under isoflurane anesthesia, using a single ligature of 3-0 silk tied around the portal vein, a calibrated constriction was performed with a 19-gauge blunt-tipped needle. The needle was then removed, leaving a calibrated constriction of the portal vein. For *in vivo* studies, animals were randomized to study the efficacy of E-VEGF-C. In cirrhotic models of portal hypertension, three groups were prepared: healthy control group, vehicle group (CCl4-V), and treated E-VEGF-C group (E-VEGF-C). Control group received olive oil 1.0 ml/kg intraperitoneal injection (i.p.) two times a week for 12 weeks and then saline 1.0 ml/kg per orally on alternate days for two weeks during the 13^th^-14^th^ week. In CCl4-V, one week after ascites formation without CCl4, RMs without VEGF-C were administered via an oral route on alternate days for 2 weeks during the 13^th^-14^th^ week. In treated E-VEGF-C group, one week after ascites formation without CCl4, E-VEGF-C was administered orally at a dose of 600μg/kg of the body weight on alternate days for 2 weeks during the 13^th^-14^th^ week. These rats were sacrificed 48 hrs. after the last dose of saline, vehicle, or E-VEGF-C, after performing the hemodynamic studies. Ascites were graded as mild, moderate, or severe based on fluid volume in intraperitoneal space. Less than 10 ml was mild, 10-30 ml was moderate, and >30 ml was severe. In non-cirrhotic model of portal hypertension, PPVL rats were prepared and randomized into two groups: (1) PPVL vehicle and PPVL+E-VEGF-C. In the PPVL+vehicle group, the RMs was administered orally on alternate days for 2 weeks after one week of surgery. In PPVL+E-VEGF-C group, E-VEGF-C was orally administered at a dose of 600 μg/kg of body weight on alternate days for 2 weeks, starting one week after surgery. The plasma volume was measured using Evans Blue dye.

### Assessment of Hepatic and Systemic Hemodynamic Parameters

Rats were anesthetized with ketamine hydrochloride (60 mg/kg) and midazolam (3mg/kg) i.p., fastened to surgical board and maintained at constant temperature of 37°C ± 0.5°C. Tracheostomy and endotracheal cannulation (PE-240 catheter; Portex, Minneapolis, MN, USA) were performed to maintain adequate respiration during anesthesia. The femoral artery and ileocolic vein were cannulated with PE-50 catheters to measure mean arterial pressure (mmHg) and portal pressure (PP) (mmHg), respectively. Non-constrictive perivascular ultrasonic transit time flow probe (2PR, 2-mm diameter; Transonic Systems Inc., Ithaca, NY, USA) was placed around the portal vein as close as possible to the liver to measure portal blood flow (PBF) (mL/min). Intrahepatic vascular resistance (IHR) (mmHg·mL·min^−1^·g^−1^) was calculated as PP/PBF. Blood pressure and flow were registered on a multichannel, computer-based recorder using Chart, version 5.0.1, for Windows software (PowerLab; AD Instruments). Hemodynamic data were collected after a 20-minute stabilization period

### Analysis of Lymphatic Transport using Tracer dye BODIPY FL-C16

To evaluate lymphatic drainage in the mLVs, a long-chain fluorescent fatty acid (BODIPY, Thermo fisher), known as lymphatic tracer, was administered orally with olive oil in 1:10 ratio with final volume of 200ul. After 2hrs., mesentery tissue was isolated and optically cleared using methyl salicylate. Images of mesenteric lymphatic drainage were obtained using confocal microscopy at 10X (Leica SP8). Six fields from each slide were randomly selected, and photographs were taken. Lymphatic drainage was quantified by measuring BODIPY fluorescence intensity inside the LVs using ImageJ. LVs leakage was quantified by measuring BODIPY fluorescence intensity in vicinity of LVs using ImageJ. Diameter of the mLVs were calculated using ImageJ diameter plugin.

### Analysis of Plasma Volume

Plasma volume was measured using Evans blue dye as previously described [6]. Briefly, 0.2 ml of Evans Blue (Himedia labs) solution (3 mg) was injected through a jugular vein catheter, followed by saline to clear the dye from the catheter. After five, 1 ml of blood was withdrawn from the femoral artery catheter. The plasma aliquot was diluted ten times in distilled water, and the absorbance was read at 600 nm using a spectrophotometer. Plasma volume (ml) was calculated using the following formula:

Plasma volume (ml) = Absorbance of standard/Absorbance of sample × 10, where the standard was 3 mg of Evans blue dye in 10 ml of plasma diluted by a factor of 10

### Computed tomography analysis

Computed tomography (CT) was performed 48 hrs. after the last dose of E-VEGF-C to visualize ascites. The animals were evaluated using Somatom Definition AS plus 128 Acquisition 384 slice reconstruction (Siemens Healthineers, Forchheim, Germany).

### Assessment of Bacterial Translocation

GFP-labelled Salmonella typhimurium was cultured in Luria Broth with ampicillin, and 10^9^ cells were orally gavaged in rats for 48 hrs. All experiments were performed under sterile conditions. Rats were anesthetized with ketamine hydrochloride (60 mg/kg) and midazolam (3mg/kg) i.p., shaved, and the skin was disinfected with alcohol. Subsequently, after midline laparotomy, MLNs were dissected, removed, and weighed in sterile condition. Tissue samples of liver, spleen, and lung were also removed and weighed. All specimens were diluted in phosphate-buffered saline (100uL per 100mg) and homogenized, and suspension was cultured on Luria broth agar with ampicillin and observed after 24 hrs. Presence of GFP-positive bacteria using UV transilluminator was considered evidence of BT to different organs. To test the translocation of bacteria in blood, 5 ml of blood was also collected in sterile condition and added to blood culture bottle, and incubated at 37 C. After 1 hr. of incubation, 100ul of blood was collected from culture bottle and spread on LB agar plate and incubated at 37 C.

### Statistical Analysis

Continuous variables are expressed as either mean<u>±</u>standard deviation for continuous distribution or as median values for skewed distribution. Continuous variables were compared between the two groups using an unpaired two-tailed Student’s t-test or Mann-Whitney U test. Variables greater than 2 were compared using one-way ANOVA followed by a post-hoc Tukey test. Bar diagrams with various data points, dot plots, and box whisker plots were plotted using GraphPad Prism (version 8.0.1.GraphPad Software, San Diego, CA, USA), and statistical analysis was performed using GraphPad Prism. Statistical significance was set at p<0.05.

## Results

### Mesenteric lymphatic vessels are dilated in experimental liver cirrhosis

To first evaluate mesenteric LVs (mLVs) in liver cirrhosis, we performed immunohistochemical staining of mesentery sections in control rats and experimental models of liver cirrhosis with antibodies recognizing Podoplanin (Pdpn), widely accepted marker of lymphatic vasculature. Control rats showed sporadic, thin LVs in vicinity of blood vessels, whereas cirrhotic rat mesentery contained numerous, readily detectable, and dilated LVs compared to control (**Figure 1A-C**). Quantitative analysis of same section revealed that the total number of Pdpn+ LVs per field in CCl4 rats was significantly higher with increased diameter compared to control (**Figure 1B,C** p<0.05). The mRNA level of LVs markers, Prox1, and LyVE1, were also enhanced in mesentery of cirrhotic rats compared to control (**Figure 1D**, p<0.05). Also, CCL21, COX2, and eNOS mRNA level was upregulated, reflecting an inflamed gut. VEGF-C expression was upregulated in CCl4 rat mesentery, suggesting inflammation-induced lymphangiogenesis (p>0.05, **Figure 1D**). VEGF-C/VEGFR3 axis is pronounced in intestinal lymphangiogenesis and inflammation-induced lymphangiogenesis[16], therefore, we checked the expression of VEGFR3 by immunofluorescence staining in mesentery sections. The protein expression of VEGFR3 was significantly increased in cirrhotic rats compared to that seen in control (**Figure 1E,F** p<0.01).

### Development of Nanoengineered VEGF-C for targeted delivery to Mesenteric Lymphatic Vessels

Dilation and lymph transport failure of the mLVs in experimental cirrhosis with ascites were previously reported[12], therefore, we hypothesized that enhancing the number of new functional LVs in mesentery by treatment with lymphangiogenic factors, VEGF-C, may improve lymphatic drainage in cirrhotic rats. To this end, we developed an engineered VEGF-C (E-VEGF-C) nanoformulation to enhance LVs uptake in the intestine after oral delivery (**Supp-Figure 1**). Human recombinant VEGF-C protein (rhVEGF-C-Cys156Ser), which specifically binds to VEGFR3 homodimer present on LyECs, was encapsulated inside the lipocarriers prepared with distearoyl-rac-glycerol-PEG2K (**Figure 2A**). In dynamic light scattering (DLS) analysis, the z-average/mean particle size (MPS) of E-VEGF-C was found to be 134.8 ± 0.47 nm with a polydispersity index (PDI) value of 0.126 ± 0.01 (**Figure 2B**). The zeta potential (ZP) and pH value of the E-VEGF-C was -21.9 ± 1.24 mV and 6.369 Â ± 0.004, respectively (**Figure 2C**). Stability study up to one month indicated no drastic change in the physicochemical properties of E-VEGF-C nanoformulation **(Supp-Table 1)**. The surface morphology and size of E-VEGF-C were studied using field emission scanning electron microscope (FE-SEM), and field emission-transmission electron microscopy (FE-TEM) indicated that particles were spherical and smooth in appearance and uniformly distributed (**Figure 2D,E**). Atomic force microscopy (AFM) micrographs provided two-dimensional and three-dimensional images of E-VEGF-C surface morphology and showed that E-VEGF-C was compact, smooth, and spherical in shape (**Figure 2F**). Average diameter of individual particles was 16nm with encapsulation efficiency of 93.28±1.85%. The release profile of E-VEGF-C showed an initial burst release of 31.95±1.52% VEGF-C observed at 2 hrs. post mixing, followed by an increase in VEGF-C release of 84.66±1.82% at 4 hrs., which then gradually decreased at other post-mixing time points (**Figure 2G)**. After the *in vitro* release study, R^2^ values obtained from different kinetic models listed in **Supp-Table 2** suggested the VEGF-C release from RMs showed zero order kinetic and, therefore, independent of concentration. To investigate the internalization of E-VEGF-C *in vitro*, we isolated LyECs from rat mesentery and mesenteric lymph nodes (MLN) using fluorescence-activated cell sorting (FACS) **(Supp-Figure 2A,B)** and incubated with coumarin-6-labelled E-VEGF-C. Fluorescence microscopy revealed the efficient internalization of E-VEGF-C by LyECs at 4hrs. **(Figure 2H,I)**. Next, to ensure the specificity of E-VEGF-C delivery *in vivo*, tissue biodistribution studies of coumarin-tagged E-VEGF-C were performed 2hrs. post its oral administration using spectrofluorimetry and fluorescence microscopy **(Figure 2J)**. A weak fluorescence signal was observed in all tissues in control and CCl4 rats **(Supp-Figure 2C)**, with an intense signal observed in mesentery of both **(Figure 2K,L)**. CCl4 rats also showed increased human VEGF-C levels in mesenteric and duodenal tissues compared to control **(Figure 2M, P< 0.05)**. In the serum of CCl4 animals, levels of VEGF-C exhibited a biphasic peak; the first peak appeared at about 10 min, and second peak appeared at approximately 5hrs. **(Figure 2N)**. There were no adverse effects or mortality after E-VEGF-C treatment in either control or CCl4 rats.

**Figure 2:**
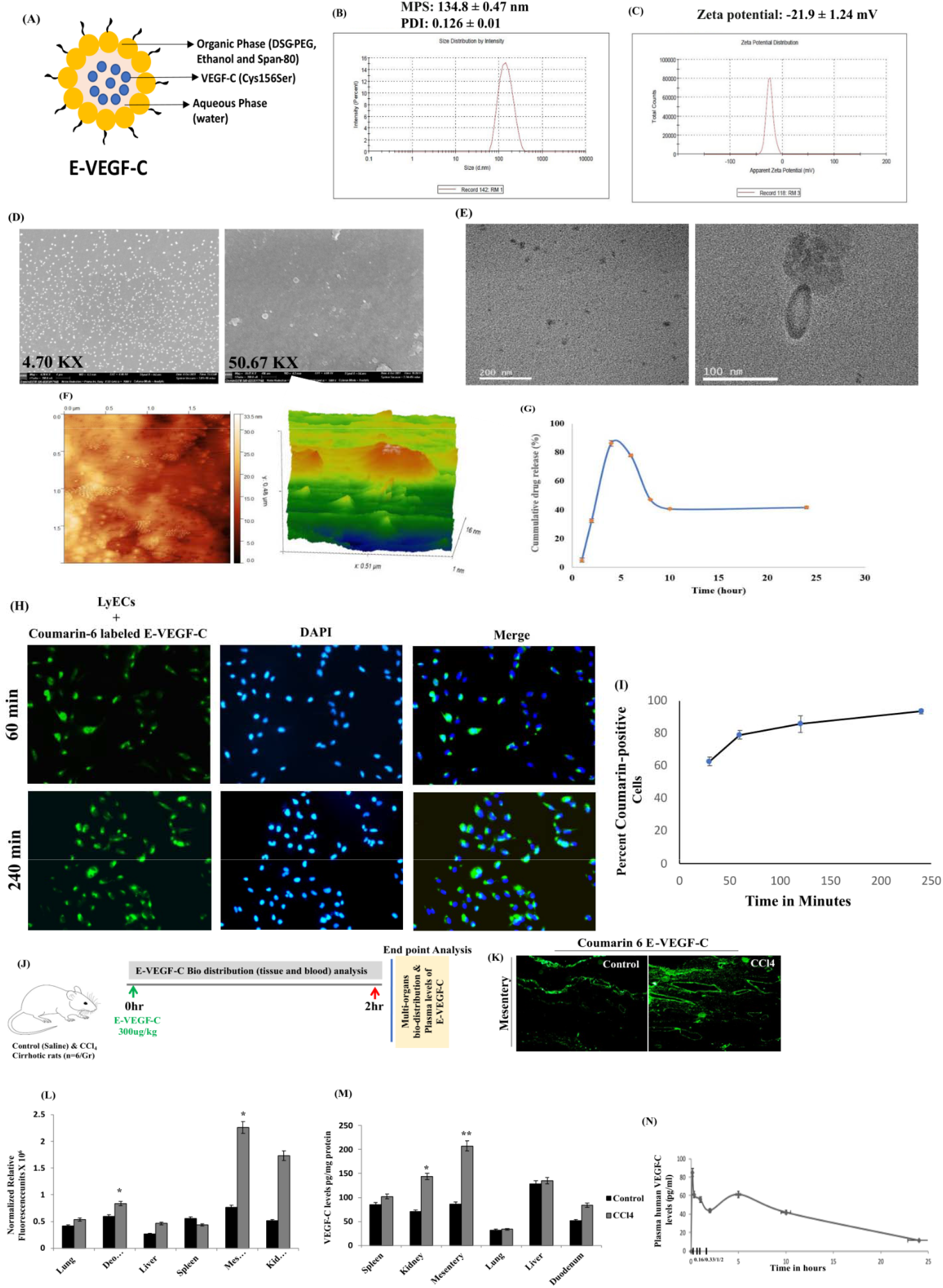
Formation, characterization, uptake and biodistribution of E-VEGF-C. **(A)**Schematic representation of VEGF-C-engineered stealth nano-lipocarriers(E-VEGF-C). Histograms showing **(B)**Mean Particle Size(MPS), Polydispersity Index(PDI), and **(C)**zeta potential of E-VEGF-C. **(D)**Field emission-scanning electron microscope(FE-SEM) and of E-VEGF-C. Surface morphology and size of E-VEGF-C were visualized using FE-SEM at 4.70KX and 50.67KX magnification. **(E)**Field emission-transmission electron microscopy(FE-TEM) analysis of E-VEGF-C The morphology and size of E-VEGF-C were examined using FE-TEM, indicating a spherical shape. **(F)**Atomic force microscopy (AFM) micrographs provide 2D and 3D images of E-VEGF-C surface morphology as compact, smooth, and spherical in shape. The average diameter of an individual particle of the prepared E-VEGF-C was 16 nm. (**G)***In vitro* release profile of E-VEGF-C in PBS at pH 7.4. Initial burst release of VEGF-C at 2hrs and 4hrs post mixing is 31.95±1.52% and 84.66±1.82% respectively. **(H)**Primary lymphatic endothelial cells(LyECs) were cultured and incubated with coumarin-6 labelled E-VEGF-C and observed at different time points. Scale bars:100μm. **(I)**Line graph of coumarin positive LyECs depicting percentage of E-VEGF-C uptake at different time points. **(J)**Schema of *in vivo* biodistribution studies. 300ug/kg of coumarin labelled E-VEGF-C was given orally in control and CCl4 rats. n=6 each. All tissues were collected after 2hrs and measured in fluorimeter. **(K)**Representative fluorescence images of mesentery section of control and CCl4-V rats showing localization of coumarin tagged E-VEGF-C in gut. Scale bars:200μm. **(L)**Quantitative fluorescence analysis in different tissue of control and CCl4 rats using fluorimeter. Data expressed as mean±SD. n=6 each group. **(M)**ELISA of human VEGF-C (pg/mg of total protein) in different tissue of control and CCl4 rats. n=3 each. *P=*0.01-0.001. **(N)**Levels of human VEGF-C (pg/ml) in control and CCl4-V rat plasma samples at indicated time points were measured using ELISA. n=3 each. Data expressed as mean±SD. ‘*’represents p< 0.05 and ‘**’represents p< 0.01

### Mesenteric Lymphatic Vessel proliferation by E-VEGF-C

Next, we studied the therapeutic effects of E-VEGF-C in animal models of liver cirrhosis. Rats were intraperitoneally injected with CCl4 for 12 weeks or till ascites formation. One week after ascites formation, 600μg/kg of E-VEGF-C was administered orally on consecutive days for up to 2 weeks (**Figure 3A**). The animals were sacrificed at 48hrs. after last dose of E-VEGF-C. Immunohistochemical analysis of mesentery showed increased numbers of Pdpn+ mLVs with reduced diameter in E-VEGF-C rats compared to CCl4-V (p<0.05, **Figure 3B-D**). Along with this, marked lymphatic vessel proliferation was observed in MLNs of E-VEGF-C rats compared to CCl4-V and control (p<0.05, **Figure 3E,F**). Mesenteric tissues also revealed a reduction in inflammation in E-VEGF-C rats compared to that seen in CCl4-V, which correlated with reduction in mRNA expression of inflammatory markers eNOS and iNOS (**Supp-Figure 3A,B**). Next, we analyzed the protein expression of VEGF-C in mesentery tissue of different study groups. Results demonstrated a significant increase in human VEGF-C protein in mesentery with MLN tissue extracts of E-VEGF-C treated rats compared to vehicle (p<0.01, **Figure 3G,H**). Since VEGF-C also participates in angiogenesis, therefore we evaluated whether E-VEGF-C had any effects on angiogenesis in mesentery. We observed no significant difference in number of CD31+ blood vessels between CCl4-V and E-VEGF-C rats (p>0.05, **Supp-Figure 3C,D**).

**Figure 3:**
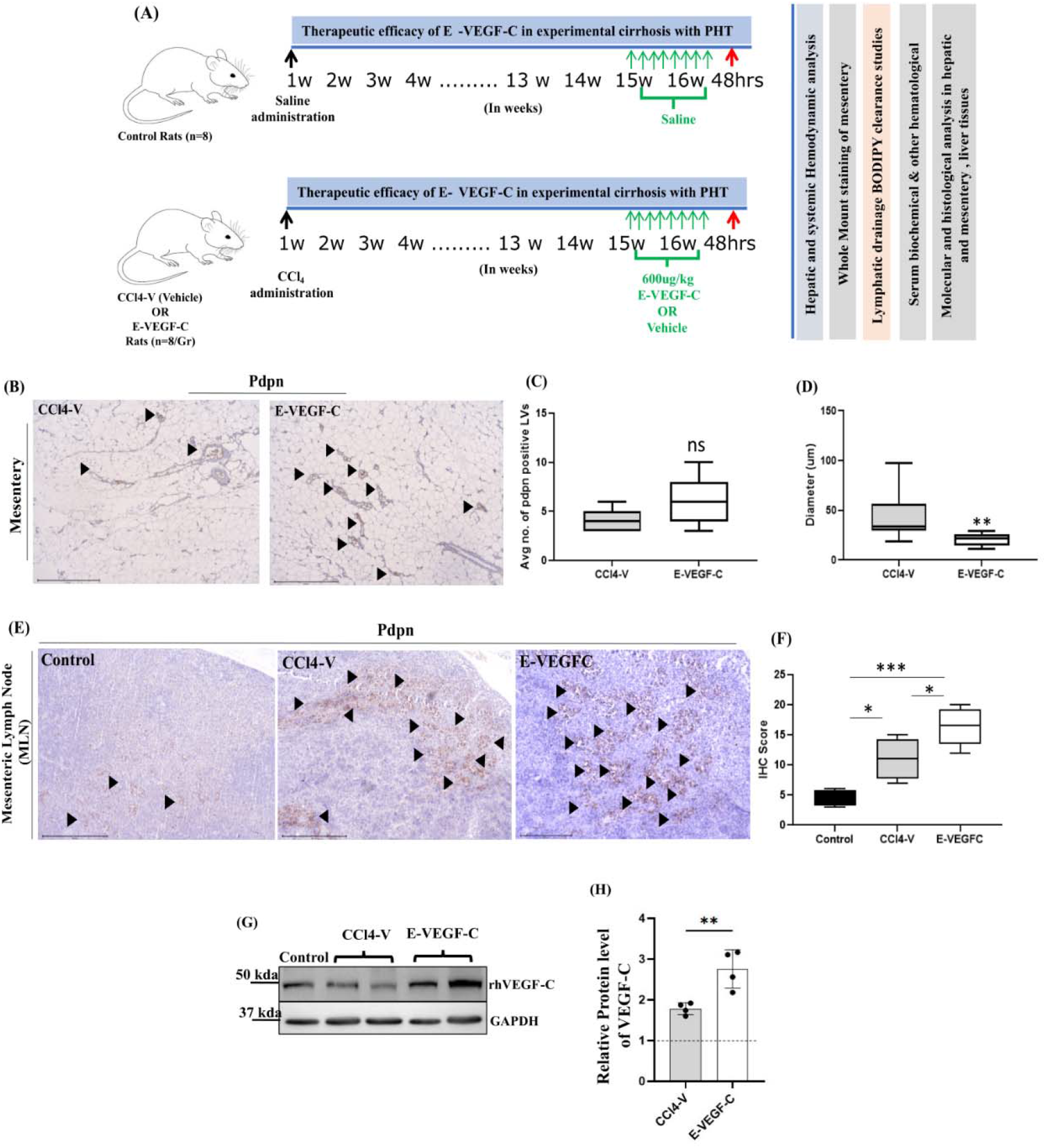
Effect of E-VEGF-C treatment on mesenteric lymphatic vessels (mLVs) in cirrhotic rat model of portal hypertension (PHT). (**A)**Schema of *in vivo* studies. CCl4 rats were treated with E-VEGF-C 600ug/kg on alternate days for 2 weeks after ascites formation. n=8 each group. **(B)**Immunohistochemistry staining for pdpn antibody in the mesenteric sections of CCl4-V and E-VEGF-C treated rat. Scale bars:500μm. Arrow indicated at pdpn+ LVs. (**C)**Number of pdpn+ mLVs were quantified using ImageJ. n=5 or 6 rats per group. Data expressed as mean± SD. Unpaired two-tailed T-test were performed. *P* > 0.05. (**D)**Diameter of pdpn+ mLVs from the same sections was measured using ImageJ diameter plugin. n=5 or 6 rats each group. Data expressed as mean±SD. Unpaired two-tailed T-test were performed. *P=*0.0073. **(E)**Immunohistochemical staining for pdpn antibody was performed on mesenteric lymph node (MLN) section of all study groups. **(F)**Brown areas were quantified as IHC scores using ImageJ. n=5 each. Data expressed as mean±SD. One-way ANOVA with Tukey’s post-hoc test was performed. *P=*0.0197 for control vs CCl4-V and *P=*0.0346 for CCl4-V vs E-VEGF-C. (**G)**Expression of human VEGF-C protein was measured using western blotting in all study groups. (**H)**Quantitative analysis of western blot is represented in bar graph. Dotted line represents control. n=4 each. Data expressed as mean±SD. ‘*’represents p< 0.05, ‘**’represents p< 0.01, and ‘***’represents p< 0.001.*represent comparison with control.

### Improved Drainage of Mesenteric Lymphatic Vessels by E-VEGF-C

To investigate the effect of E-VEGFC on LVs patterning, whole-mount immunostaining of mesentery was performed using pdpn antibody. In control rats, thin LVs of diameter ranging from 50 to 80μm were observed, which were increased to 100 to 150μm in CCl4-V rats. In E-VEGF-C treated rats, sprouting of new LVs from the existing one was clearly observed with reduced diameter ranging from 60 to 100μm compared to the CCl4-V rats (**Figure 4A,B**). Branching points of mLVs close to intestine were also increased in E-VEGF-C rats in comparison to CCL4-V (**Figure 4C,D**). To assess the functionality of LVs, we gavaged rats with BODIPY FL-C16 and analyzed drainage and leakage of mLVs after 2hrs. (**Figure 4E)**. CCl4 rats displayed increased BODIPY fluorescence inside mLVs with an increased diameter compared to the control rat’s mLVs, which represents accumulation of dye and, hence, incomplete drainage. On the other hand, E-VEGF-C-treated rats had significantly reduced fluorescence inside the mLVs with decreased diameter (p<0.001, **Figure 4F-H**). Dilated LVs cause leakage of the lymph and accumulation in tissue spaces. To assess leakage from mLVs, fluorescence outside mLVs was quantified. We observed increased fluorescence outside the LVs in CCl4 rats compared to control, which was significantly reduced in E-VEGF-C rats (p<0.001, **Figure 4I**).

**Figure 4:**
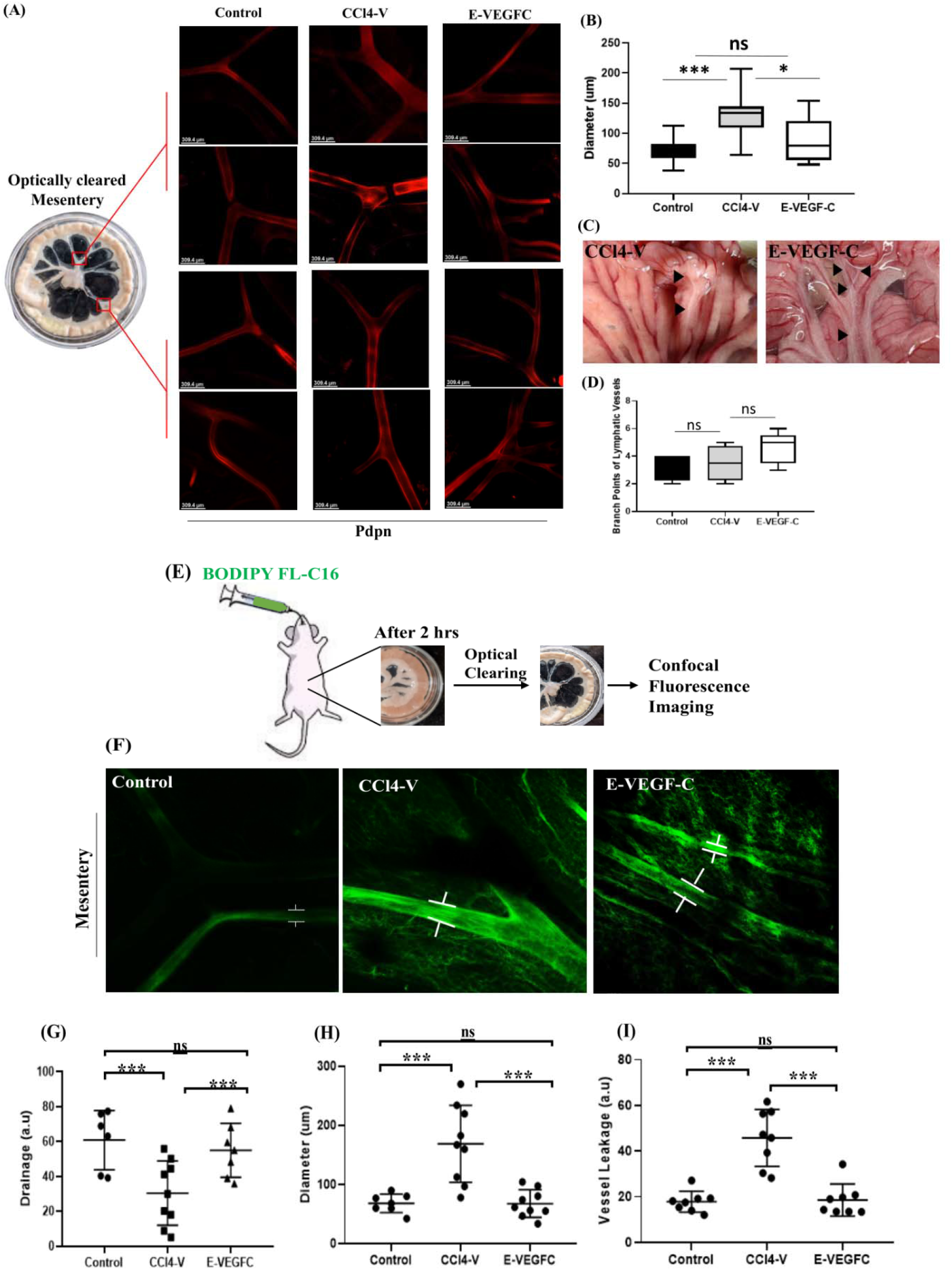
Effect of E-VEGF-C on proliferation and drainage of mLVs. (**A)**Whole-mount immunostaining of mesentery for visualization of mLVs in all study groups was performed with pdpn antibody. The upper 2 panels represent mesenteric collecting LVs, and the lower 2 panels represent lymphatic capillaries entering the intestine. Scale bar:309.4μm. (**B)**Diameter of collecting mLVs were measured from same section using ImageJ diameter plugin. n=3 or 4 rats each group. Data expressed as mean± SD. One-Way ANOVA with post-hoc Tukey’s test was performed. *P*<0.0001 for control vs. CCl4-V and *P=*0.0183 for CCl4-V vs. E-VEGF-C (**C)**Mesentery tissue of CCl4-V and E-VEGF-C rats. Arrowheads indicated proliferation and branching of mLVs in mesentery of E-VEGF-C treated rat. (**D)**Quantitative analysis of branching points in mesentery in all study groups. n=3-4 rats each group. Data expressed as mean±SD. One-Way ANOVA with post-hoc Tukey’s test was performed. *P*>0.05 for each comparison. **(E)**Workflow for visualization of mLVs by oral gavaging of BODIPY FL-C16. (**F)**Corresponding images of mLVs after BODIPY FL-C16 administration. Scale bar:309.1μm Representative graphs for characterization of LVs. Mean of three points from each field was taken. **(G)**Diameter of LVs was measured in μm using ImageJ diameter-plugins. (**H)**Drainage of LVs was measured by fluorescence inside the vessels using ImageJ by quantifying mean intensity of BODIPY. 100-mean intensity percentage were plotted. (**I)**Quantification of lymph leakage from vessels by measuring mean fluorescence intensity of BODIPY in extraluminal space. n=4 or 5 rats each group. Data expressed as mean±SD. ‘***’represents p<0.001, and ‘*’represents p<0.05.

### E-VEGF-C reduces Ascites and ameliorates Portal Pressure in Cirrhotic and Non-cirrhotic Portal Hypertensive Rats

We next probed whether an improvement in lymphatic drainage, with proliferation of mLVs in E-VEGF-C treated rats had any effect on ascitic fluid volume. CT scan analysis of all study groups was performed to observe ascites in the peritoneal cavity. No ascites was observed in the control group, whereas all CCl4-V rats displayed severe ascites **(Figure 5A)**. E-VEGF-C rats displayed marked reduction in ascites volume as compared to CCl4-V (P< 0.001, **Figure 6A,B, Supp-Table 3**). Along with ascites reduction, evans blue staining showed a significant increase in plasma volume in E-VEGF-C group compared with CCl4-V(P< 0.05, **Figure 5C**).

**Figure 5:**
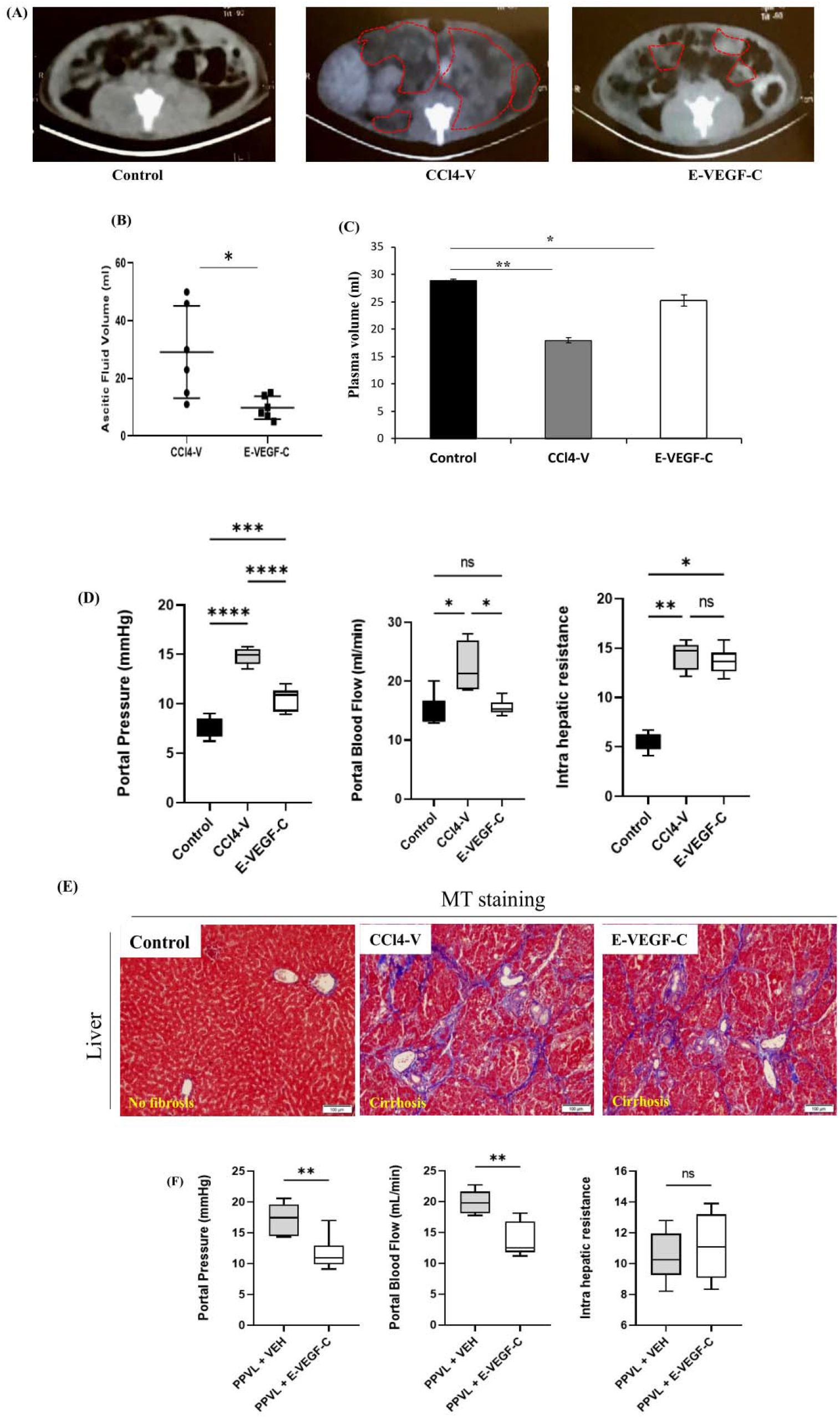
Effect of E-VEGF-C treatment on ascitic fluid volume and hemodynamic parameters in CCl4 and Partial Portal Vein Ligation (PPVL) rat model. **(A)**Representative CT scan slices of all study groups showing abdominal cavity. Regions of interest are marked with red dotted outline and correspond to the zones with fluid accumulation. **(B)**Dot plots showing ascitic fluid volume(ml) in CCl4-V and E-VEGF-C treated rats. n=6 each group. Data expressed as mean±SD. Unpaired two-tailed t-test were performed, *P=*0.0168. (**C)**Histograms showing plasma volumes(ml) in all study groups measured using the Evans Blue dye dilution technique. n=4 each. (**D)**Bar Diagrams showing hepatic hemodynamic parameters, Portal pressure(PP), Portal Blood Flow(PBF), and Intrahepatic resistance(IHR) in study groups n=6 each. Data expressed as mean±SD. One-Way ANOVA with post-hoc Tukey’s test was performed. *P*<0.05 **(E)**Masson Trichrome(MT) stained images (10x) of liver tissues in different animal groups. Liver fibrosis was assessed using the Laennec fibrosis scoring system. (**F)**Bar Diagrams showing hepatic hemodynamic parameters Portal pressure(PP), Portal Blood Flow(PBF), and Intrahepatic resistance(IHR) in control, PPVL vehicle and PPVL+E-VEGF-C rats. n=5 each Data expressed as mean±SD. Unpaired two-tailed t-test were performed. ‘*’represents p< 0.05 and ‘**’represents p< 0.01, and ‘***’represents p< 0.001. *Represent comparison with control.

**Figure 6:**
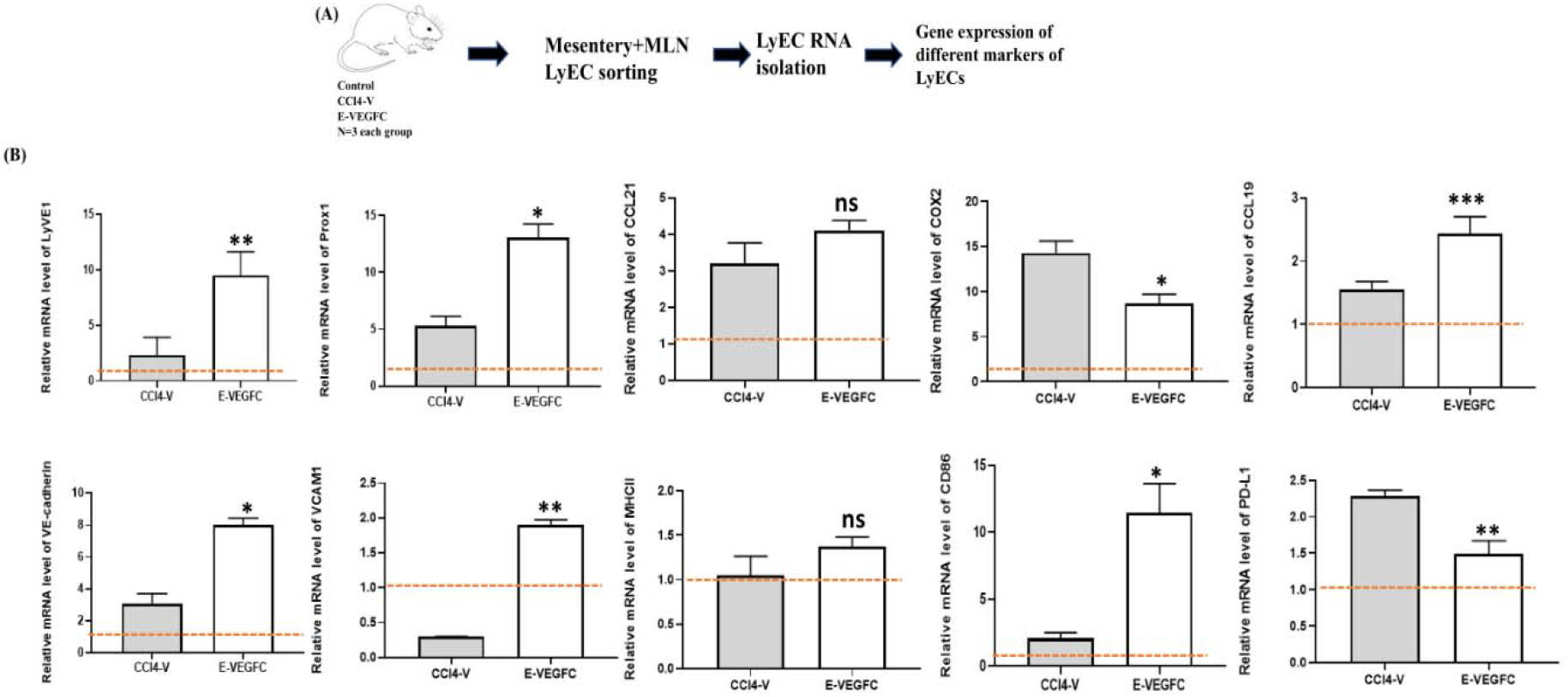
Effect of E-VEGF-C on gene expression profiling of lymphatic endothelial cells (LyECs). **(A)**Schema of workflow. LyECs were isolated from mesentery and MLNs of all study groups using FACS. RNA isolation and qRT-PCR analysis were done for different marker genes of LyECs. **(B)**Relative gene expression of LyVE1, Prox1, VCAM1, VE-Cad, MHCII, CD86, CCL21, and COX2 genes in CCl4-V and E-VEGF-C treated rats were plotted. Dotted lines represent control. n= 3 each. Data expressed as mean±SD. One-Way ANOVA with post-hoc Tukey’s test was performed. ‘**’represents p<0.01,’*’represents p<0.05. *Represent comparison with CCl4-V.

Next, we analyzed whether this reduction in ascitic fluid volume after E-VEGF-C treatment was also associated with liver physiology and pathological changes. Intraperitoneal CCl4 for 12-weeks markedly increased portal pressure (PP) in rats compared to that in control (p<0.001, **Figure 5D**). However, E-VEGF-C treatment significantly attenuated PP compared to vehicle (p<0.001, **Figure 5D**). To ascertain whether the observed reduction in PP was due to change in portal blood flow (PBF) or intrahepatic resistance (IHR), we also monitored these parameters. E-VEGF-C treatment significantly decreased PBF, in turn increasing the mean arterial pressure (MAP) as compared to CCl4-V(p<0.05, **Figure 5D, Supp-Figure 4**). However, IHR in E-VEGF-C-treated rats was similar to that in CCl4-V (p>0.05, **Figure 5D**).

Compared to control, liver weights in E-VEGF-C and CCl4-V rats were also similar (**Supp-Figure 4**). We next evaluated changes in histological and biochemical parameters of liver in cirrhotic rats. Masson’s trichrome staining of liver tissues revealed that both CCl4-V and E-VEGF-C rats showed Ishak fibrosis scores of 5/6 or 6/6, indicating complete cirrhosis (**Figure 5E**). CCl4 resulted in a significant increase in serum ALT levels and decreased albumin levels compared to control (**Supp-Table 3**). There was no significant improvement in serum albumin and ALT levels in E-VEGF-C-treated rats compared to CCl4-V. Kidney functions were normal in all study groups.

We also investigated whether E-VEGF-C treatment improved PP irrespective of liver cirrhosis by measuring hepatic hemodynamics in non-cirrhotic portal hypertensive (PPVL) rats. Histology of these animals displayed portal inflammation in liver but no significant fibrosis (**Supp-Figure 5A,B**). Hemodynamic analysis revealed significant reduction in PP and PBF of E-VEGF-C treated PPVL rats compared with vehicle (p<0.01). Along with this, E-VEGF-C-treated PPVL rats showed an increase in number of VEGFR3+ mLVs and VEGF-C protein expression with reduced inflammation in mesentery compared to PPVL-vehicle. E-VEGF-C-treatment also reduces the mRNA expression of iNOS and eNOS in mesentery compared to the vehicle (p<0.05, **Supp-Figure 5C-F**).

### E-VEGF-C treatment modulates gene expression in lymphatic endothelial cells of mesentery and lymph nodes

To gain mechanistic insights into VEGF-C-induced lymphangiogenesis in mLVs, LyECs from mesentery and MLNs were isolated using FACS and expression of relevant genes was examined (**Figure 6A**). The mRNA level of differentiation and proliferation markers of LyECs, i.e., Prox1 and LyVE1 were upregulated in LyECs of E-VEGF-C treated rats compared to vehicle (p<0.05, **Figure 6B**). Collecting LVs generally present in mesentery, prevents any leakage of lymph by zippering between LyECs using VE-cadherin and VCAM-1. mRNA levels of VCAM1 decreases in LyECs of CCL4-V rats, indicating leaky mLVs causing more inflammation. No significant change was observed in VE-cadherin in CCl4-V rats compared to control. Treatment with E-VEGF-C led to increased expression of both VCAM1 and VE-cadherin in LyECs correlating with decreased leakage of lymph from mLVs (p<0.05). LyECs also act as APCs, present Ag to T cells for tolerance or induction of immune response. Costimulatory marker, CD86, was also significantly upregulated in LyECs of treated rats. In contrast, no significant change was observed in MHC-II levels in vehicle vs. treated group (p>0.05). The inflammatory marker COX2 was markedly increased in LyECs of CCl4-V rats suggesting inflamed lymphatic endothelium, which further reduced in LyECs of treated group (p<0.05). CCL21 and CCL19 chemokine mRNA level is found to be increased in LyECs of E-VEGF-C treated rats compared to vehicle resulting in increased immune cell trafficking to MLNs (p<0.05). PD-L1 was increased in LyECs of CCl4-V rats which further reduced in treated group (p<0.01).

### E-VEGF-C increases trafficking of immune cells and clearance of bacteria load in mesenteric lymph node

VEGF-C is known to increase immune cell trafficking to the lymph nodes[17,23]. Therefore, to study the effect of E-VEGF-C treatment on immune cell trafficking to MLNs, cells were isolated from MLN of all study groups and labeled with antibodies for T cell subsets and DCs for quantification using flow cytometry (**Supp Figure 6A,B**). Total T cell, Th and Tc cell population in MLNs did not change significantly in control and CCl4-V as previously reported[24]. Treatment with E-VEGF-C also led to no change in the above-mentioned population. We further quantified recently activated T cell with CD134 and regulatory T cell with CD25 expression. No significant change was observed in CD134 expression in varied groups (**Figure 7A,B)** but we observe a significant decrease in CD8 Treg cells after E-VEGF-C treatment in comparison to CCl4-V (**Figure 7C-D**). DCs, along with the expression of T cell activation coreceptor CD80, were increased after treatment with E-VEGF-C compared to that observed in the vehicle (p<0.05, **Figure 7E**).

**Figure 7:**
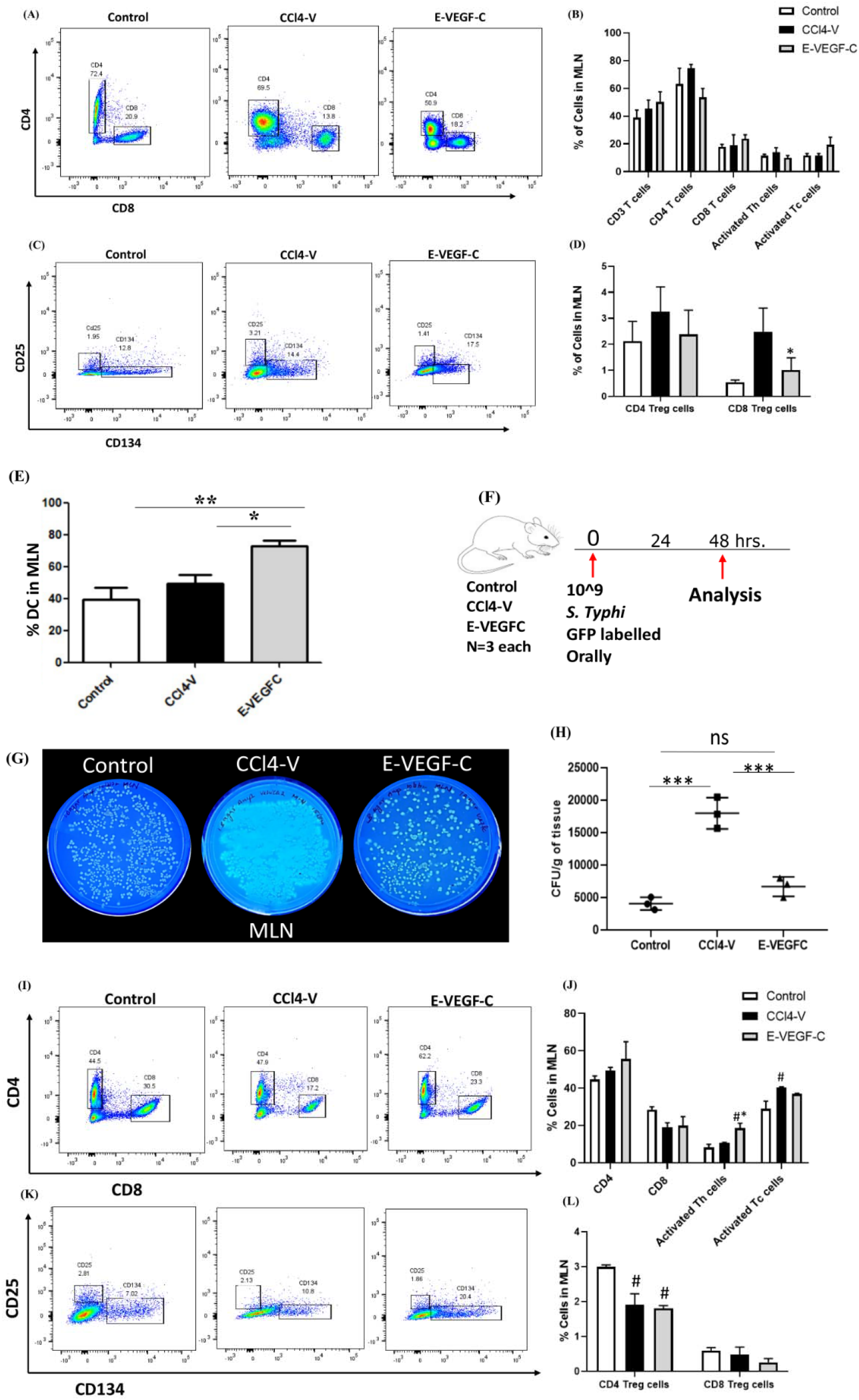
Effect of E-VEGF-C on priming of immune cells and clearance of bacterial load. (**A)**Dot Plots of T cells subsets in MLN of all study groups. **(B)**Percentage-population of CD3 T cells, CD4 helper T cells, CD8 cytotoxic T cells, CD134+ recently activated helper and cytotoxic T cells. **(C)**Dot plots of CD25+ regulatory helper T cells in all study groups. **(D)**Percentage-population of T regulatory cells positive for CD3, CD4/CD8, and CD25. (**E)**Percentage-population of dendritic cells(DCs) positive for CD11c, CD103, and CD80. n=5 each. Data expressed as mean±SD. One-Way ANOVA with post-hoc Tukey’s test was performed. *P*<0.05. **(F)**Schema of workflow, 10^9^ GFP+ bacteria were given to rats orally, and tissue were collected in sterile condition after 48hrs of gavage. 100mg of MLN tissue were used for bacterial load quantification. Cells were isolated from MLNs for quantification using flow cytometry. (**G)**GFP+ *S. typhimurium* colonies in 100mg of MLN tissue extract of each group visualized using UV transilluminator. (**H)**Quantitative analysis of CFU/gm of MLN tissue for GFP+ *S. typhimurium*. n=3 each. Data expressed as mean±SD. One-Way ANOVA with post-hoc Tukey’s test was performed. *P*<0.001. **(I)**Dot Plots of T cells subsets in cells isolated from MLNs of all study groups after bacterial challenge. **(J)** Percentage-population of CD4 helper T cells, CD8 cytotoxic T cells, CD134+ recently activated helper, and cytotoxic T cells. **(K)**Dot plots of CD25+ regulatory helper T cells. **(L)**Percentage-population of T regulatory cells positive for CD3, CD4/CD8, and CD25. n=4 each. Data expressed as mean±SD. One-Way ANOVA with post-hoc Tukey’s test was performed.’#’ represents comparison with control.’*’ represent a comparison with CCl4-V.’***’represents p<0.001,’**’represents p<0.01, and ‘*’represents p<0.05.’***#***’represents p<0.05.

Though we observed no significant changes in T cell subsets in MLNs of either CCl4-V or E-VEGF-C, we wanted to look for any change in immune response in the presence of antigen challenge, therefore, we performed a few experiments with live bacteria[25]. To analyze the effect of activated T cells on bacterial clearance in MLNs after E-VEGF-C treatment, we gavaged rats with GFP-labelled *Salmonella typhimurium* (**Figure 7F**). MLNs, liver, spleen, lung, and blood were collected after 48hrs. of incubation. In control rats, GFP-labelled bacteria were found only in MLNs, whereas in CCl4-V, GFP-labelled bacteria were present in all tissue collected, including blood (**Figure 7G, Supp-Figure 6C**). Interestingly, in E-VEGF-C treated rats, bacterial translocation was confined only to MLNs with reduced live bacteria than CCl4-V (**Figure 7G,H**), suggesting clearance of bacterial load. We also quantified T cell subsets in MLNs and found significant increase in recently activated Th cells in MLNs of E-VEGF-C treated rats compared to CCl4-V, which indicate active immune response in MLNs in the presence of antigen (**Figure 7I-L**). We also assessed the levels of TNF-α and endotoxins in ascitic fluid and serum and observed no changes in serum endotoxins and TNF-α levels in the E-VEGF-C treated rats compared to CCl4-V. However, ascitic endotoxins levels were reduced in E-VEGF-C treated rats **(Supp-Figure 7A-D)**.

## Discussion

In this study, we investigated morphological and molecular alterations of mLVs and MLNs in experimental cirrhosis and portal hypertension and reported significantly increased number of dilated pdpn+ and VEGFR3+ leaky mLVs with reduced drainage capacity and enhanced inflammatory markers in cirrhotic and portal hypertensive rats clearly implying gut lymphatic dysfunction similar to that observed in intestinal lymphangiectasia and gut inflammatory disorders[26,27]. Increase in mLVs density in cirrhosis may be attributed to compensatory lymphangiogenesis response to lymphatic occlusion[28]. A previous study in cirrhotic rats documented an increased expression of eNOS in mesenteric LyECs as the cause of dysfunctional lymphatics in cirrhosis and that inhibiting eNOS improved the lymphatic transport of the existing vessels by increasing the contractile activity of these vessels[12]. In our study, we focused on increasing the number of new functional lymphatic channels via VEGF-C treatment.

Due to short half-life and systemic effects of VEGF-C[18], we constructed recombinant VEGF-C molecule using RM-based nano-lipocarriers. RM nano-lipocarriers are nanoparticle-sized water-in-oil microemulsions with controlled particle sizes. A recent report illustrated the therapeutic potential of the fully human fusion protein F8-VEGF-C for targeted delivery of VEGF-C in mouse models of chronic inflammatory skin disease[29]. However, the study did not report sustained release of VEGF-C. In another study, VEGF-C mRNA was encapsulated in lipid nanoparticles for its sustained release at administration site and found to be effective in treating experimental lymphedema[30]. We used various strategies in this study to ensure targeted and sustained delivery of VEGF-C in mLVs. First, the use of RM-based lipid nanocarriers allowed VEGF-C cargo to be carried in chylomicron-sized particles in LVs after oral delivery. Encapsulation also ensured their sustained and programmable release[31]. *In vitro* uptake assays clearly showed more than 90% uptake of E-VEGF-C by mesenteric LyECs. *In vitro* release studies indicated more than 80% VEGF-C release in about 4hrs. and *in vivo* the release profile showed biphasic peak, with the initial burst coming at approximately 10 min, and the second peak at about 5 hrs. Biphasic drug release is a characteristic feature of drugs encapsulated in nano-lipocarriers[32]. In *in vivo* biodistribution studies as well, cirrhotic rats showed maximum expression of VEGF-C in mesentery, indicating the efficacy of our delivery vehicle. We did not follow any conjugation chemistry for the preparation of E-VEGF-C molecule, rather, we focused on preparation of simple solid lipid nanoparticles with an aqueous template using a modified multiple emulsification technique[33].

Treatment with E-VEGF-C markedly enhanced the expression of VEGF-C protein in the mesentery, along with a concomitant increase in the sprouting of pdpn+ and VEGFR3+LVs both in the mesentery and MLNs, suggestive of VEGF-C driven proliferation of LyECs[34]. The dilations observed in mLVs of cirrhotic rats were also attenuated with E-VEGF-C treatment. Specific genes, such as Prox1 and LyVE1, were significantly increased in the mesenteric LyECs of E-VEGF-C rats, indicating pronounced differentiation and proliferation of LyECs. Expression of inflammatory genes, such as Cox2, was decreased, while VCAM-1 and VE-cadherin, which govern cell adhesion and vessel permeability, increased in E-VEGF-C rats. The functional implications of enhanced gene expression were evident in terms of reduced inflammation, lymph leakage, and improved drainage in E-VEGF-C treated rats. This is in accordance with many previous studies showing the drainage-promoting function of VEGF-C in experimental models of bacterial skin inflammation and IBD[35,36]. An improvement in lymphatic drainage and functionality of the mLVs in cirrhotic rats was also associated with marked reduction in ascitic fluid volume in rats. Decrease in ascites was accompanied by an increase in plasma volume of treated rats, suggesting that ascites may have been reabsorbed into the circulating blood[11].

Along with a reduction in ascites, there was also a significant decrease in PP in cirrhotic portal hypertensive rats treated with E-VEGF-C, which was associated with attenuated PBF and increased MAP. However, there was no change in IHR, indicating that improvement in PBF and not hepatic resistance (fibrosis) led to an improvement in portal pressure after treatment. It has been postulated that the removal of ascites plays role in postparacentesis systemic hemodynamic changes through mechanical decompression of the splanchnic vascular bed[37]. However, it is plausible that a reduction in PP due to an increase in number of LVs and decrease in interstitial fluid pressure may also have led to decrease in ascites[11]. Intriguingly, this decrease in PP was also observed in the non-cirrhotic PPVL animals after E-VEGF-C treatment, validating favorable effects of VEGF-C on PBF and systemic hemodynamics. An improvement in PP was not associated with a significant improvement in liver pathology in the treated rats, indicating that there were no protective effects of E-VEGF-C treatment on hepatic compartment per se. We also did not observe any major changes in the serum albumin or ALT levels in the treated animals. In our study, we administered therapeutic E-VEGF-C treatment in decompensated animal models when liver cirrhosis and ascites had already been established. It would be worthwhile to evaluate the effects of E-VEGF-C on early onset of decompensation and liver pathology in compensated cirrhotic models of portal hypertension using a preventive treatment approach.

To note, we did not detect any significant changes in the density of CD31+blood-vessels in E-VEGF-C-treated cirrhotic rats, indicating that LV-specific VEGF-C(Cys156Ser) did not affect mesenteric blood-vessels and was specific only to mLVs as previously demonstrated[38]. With respect to inflammation, there was a conspicuous decrease in mesenteric tissue inflammation in the E-VEGF-C-treated rats.

VEGF-C is known to increase immune cell trafficking to the draining lymph node[14]. We did not observe any significant change in T cell recruitment in MLNs of E-VEGF-C treated animals as compared to vehicles. There was, however an increase in costimulatory markers, such as, CD86 in the LyECs of E-VEGF-C treated rats, suggesting active antigen presentation by these cells[39,40]. In cirrhosis, studies have reported an increased live bacterial translocation to MLNs, causing systemic spread of the infection and therefore, multiple organ failure due to sepsis and septic shock[41,42]. In our study, when we challenged the cirrhotic animals with live bacteria, we observed live bacteria not only in MLNs but also in other organs and blood, indicating an impaired gut immune response in the MLNs. In cirrhotic animals treated with E-VEGF-C, bacteria remained confined to the MLNs only, with a significant increase in recently activated CD4 T cells, suggesting an appropriate immune response in their MLNs. An earlier study documented that the stimulation of cardiac lymphangiogenesis with VEGF-C improved the trafficking of immune cells to the draining lymph nodes after myocardial infarction resolving inflammation[43]. Along with an improvement of local gut immune response, we also observed a decrease in endotoxins, suggesting attenuation of endotoxemia after E-VEGF-C treatment[44].

In summary, our study underscores the use of nanolipocarriers incorporating hydrophilic LV-specific VEGF-C as novel therapy for improving lymphatic drainage and gut immunity by providing an efficient exit route for ascitic fluid. Gut LVs-targeted delivery of E-VEGF-C may open new and exciting avenues for the treatment and even prevention of decompensation in cirrhosis.

## Data Availability

Data are available on request

## Author Contributions

SK, DMT, and SB conceptualize and designed the work. PJ, SNRR, IK, PR and SR collected the data. PJ, SNRR, SK, DMT, SB, and IK analyzed interpret the data. PJ, SNRR, SK and SB drafted the article. SK, DMT, SB, VN and SKS did the critical revision of article. All authors read and approved the final article.

## Acknowledgements

The authors would like to thank Department of Science and Technology (DST), Ministry of Science & Technology, Government of India (DST/NM/NT/2019/191) for the financial support and Center for Nanotechnology (CNT) and the North East Centre for Biological Sciences and Healthcare Engineering (NECBH) of IIT-Guwahati, Assam, India, for providing the facility for AFM and FE-SEM analysis.

## Notes

**Data Availability:** All the data supporting the findings of this study are available within the article and its supplementary information files and from the corresponding authors upon reasonable request.

### Competing Interest Statement

The authors have declared no competing interest.

### Author Declarations

Institutional Ethics Committee, Institute of liver and biliary Sciences (ILBS)

### Summary of Updates

modified the main text

