## Supplementary Methods for "Recombinant VEGF-C (Cys156Ser) protein restores mesenteric lymphatic drainage and improves gut immune surveillance in experimental liver cirrhosis"

**Table of Content:**

Material and methods………………………………………………………………………………..2

Supplementary figures and legends……………………………………………….............................7

Supplementary Table……………………………………………………………………………….16

References…………………………………………………………………………………………...18

**Material and Methods:**

**Preparation of Engineered VEGF-C (E-VEGF-C)**

To prepare RMs, Distearoyl-rac-glycerol-PEG2K (DSG-PEG2000) and Span-80 were dissolved in an organic solvent ethanol. Span-80 was used as the emulsifier. Double-distilled water containing VEGF-C was added dropwise to organic phase and mixed well with magnetic stirrer. The coarse dispersion was then passed through benchtop high-shear homogenizer at 20,000 pressures for three cycles to obtain nanosized translucent RMs containing VEGF-C termed as E-VEGF-C.

**Characterization of** **E-VEGF-C:**

***Mean particle size (MPS), polydispersity index (PDI) and zeta potential***

The MPS, PDI, and ZP values were evaluated using dynamic light scattering (DLS). One hundred microliters of RM sample were diluted with double distilled water up to 1 ml, in triplicate. Using a Malvern Zetasizer (Nano ZS, Malvern Instrument Ltd, Malvern, UK), the MPS, PDI, and ZP values of the diluted RMs were determined at 25 °C by placing 1 ml of sample directly into a standard quartz cuvette. A He-Ne light source at 633-nm wavelength was used as the source of the laser beam in this instrument.

***Stability study***

The RM formulation was observed over a period of one month at 4°C for the stability study. Samples were withdrawn at regular time intervals of 0, 1, 2, 3, and 4 weeks to observe the influence of storage conditions on MPS, PDI, and ZP values.

***pH measurement***

The pH of the RM formulation was measured in triplicate using a pH meter (Mettler Toledo, Greifensee, Switzerland) at room temperature.

***Field emission-scanning electron microscope (FE-SEM)***

The surface morphology and size of the RM formulations were examined using FE-SEM (GeminiSEM 500, Carl Zeiss Microscopy, GmbH, Oberkochen, Germany). Before loading the sample into the FE-SEM instrument, it was coated with gold to avoid or minimize the charging effect. Images were recorded at a voltage–2-4 kV using an in-lens detector [1].

***Atomic force microscopy (AFM)***

AFM (Innova SPM, Bruker, Germany) analysis was performed in the non-contact tapping mode to record the topographical images of the optimized RM formulation. The samples were prepared using the drop-casting method, followed by drying at room temperature. The raw data obtained from the system were processed using Gwyddion software, version 2.60.

***Field emission-transmission electron microscopy (FE-TEM)***

The morphologies and sizes of the RMs were examined using FE-TEM (JEOL, 2100F, Japan). Approximately 10 μl of the sample was dropped onto a carbon-coated copper grid and covered with collodion carbon, followed by air-drying at room temperature before measurements were taken. Images were recorded at an accelerating voltage of 90 kV, and images were recorded.

***Determination of*** ***encapsulation efficiency (EE %)***

The percentage of VEGF-C encapsulated in the formulation was quantified by enzyme-linked immunosorbent assay (ELISA) using the Quantikine^TM^ human VEGF-C ELISA kit protocol [2]. The prepared reverse micelles were placed in a rotary evaporator (IKA^®^, Werke GmbH & Co. KG, Stäüfen, Germany) to remove the solvent present in the formulation at 58º C temperature, 100 rpm, and -700 to -800 mbar pressure. The thin film obtained was hydrated using double-distilled water. One milliliter of the redispersed formulation was taken and 1 ml of ethanol was added to disrupt the self-assembled structure of the lipid layer. The samples collected in triplicate were analyzed using an ELISA kit at 450 nm with a multimode reader (SpectraMax^®^, CA, USA) according to the manufacturer’s instructions. The EE % was calculated using Equation (1).

$$EE \%=\frac{Amount of VEGF-C in formulation}{Actual amount of VEGF-Cadded in formulation}\times100\ldots.Eq.(1)$$

***In vitro VEGF-C release from reverse micelles***

The *in vitro* VEGF-C release from the RMs was analyzed by ELISA using the Quantikine^TM^ human VEGF-C ELISA kit protocol [3]. Two milliliters of reverse micelle dispersions containing 666.6 ng VEGF-C were placed in the middle of the dialysis bag. The dialysis bag containing the dispersions was then kept in 5 ml of release medium (phosphate buffer saline, PBS, pH 7.4) in a 50 ml falcon tube. The Falcon tube was then placed in an incubator shaker (REMI Sales & Engineering Ltd., Mumbai, India) at 37 °C with shaking at 75 rpm. The samples were withdrawn at different time points (1, 2, 4, 6, 8, 10, and 24 h), and pre-warmed fresh medium was added at each time point. The collected samples were stored at -20 °C for later analysis. The collected samples were analyzed according to the manufacturer’s instructions in the ELISA kit booklet using an ELISA kit at 450 nm with a multimode reader (SpectraMax^®^, CA, USA). The results are expressed as percent cumulative release ± standard deviation (SD) in triplicate.

The percent cumulative release data were fitted into different release kinetic models, such as zero-order, first-order, Korsmeyer-Peppas, Weibull, Higuchi, and Hixson-Crowell models, using KinetDS software, version 3.0. The best-fit plot was chosen based on the maximum R^2^ (coefficient of determination) value [4].

**In vivo studies**

***In Vivo Biodistribution and half-life of E-VEGF-C***

E-VEGF-C (single dose: 300μg/kg) tagged with coumarin-6 dye was administered on day 0 in healthy animals and CCL4 rats after 12 weeks. Rats were euthanized 2 hrs. after a single oral administration of E-VEGF-C for the collection of liver, intestine, mesentery, spleen, lung, and kidney tissues. The tissues were weighed and homogenized with physiological saline, and fluorescence intensity was detected at 520 nm using a spectrofluorometer. Some parts of the tissues were also observed under a fluorescence microscope for visual detection of labeled nanoparticles. The fluorescence intensity values of coumarin were normalized to those of the untreated control rats. Levels of VEGF-C were determined per mg protein of the tissues using the human VEGF-C ELISA kit (Elabsciences, Houston, USA) according to the manufacturer’s protocol.

For plasma half-life studies, after a single oral injection of E-VEGF-C, serial blood samples (300 μl) were collected from the rats in EDTA vials at 10 min, 20 min, 30 min, 1 hr, 5 hr, 10 hr, and 24 hr. After 20 min of incubation at room temperature, blood samples were centrifuged at 1200*× g* for 10 min. The plasma (100-150 μl) was collected, snap frozen, and stored at –80°C. The amount of VEGF-C in each sample was determined using a human VEGF-C ELISA kit, as previously described. Plasma VEGF-C concentration–time data were analyzed at different time points.

***Whole mount staining:***

For whole mount staining, mesentery was isolated with intestine and flushed with PBS and kept in 4% PFA for 1hr at RT. After incubation, mesentery was washed with PBS three times and immersed in PBST (PBS+ 0.03% triton X100) for 30 min at RT. Blocking was done with 3% BSA+0.01% triton for 2 hrs. at RT and incubated with primary antibody overnight on rocker. Mesentery was washed with PBS thrice and secondary antibody labelled with fluorochrome was added for 2 hrs. at RT on rocker. Washing was done with PBS thrice to remove unbound antibody. Mesentery was separated with intestine and mounted on slides with vectashield and imaging was done in Leica confocal microscope.

***In vitro* Studies**

***Isolation of Lymphatic endothelial cells from mesenteric tissue of rat and uptake of E-VEGF-C in vitro***

For the isolation of mesenteric LyECs, protocol was adapted from Ribera *et al.*, 2013 with minor moddifaction. Briefly, the rats were anesthetized with ketamine hydrochloride (60 mg/kg) and midazolam (3mg/kg) intraperitoneally. A midline incision was made to perfuse the mesentery with saline by inserting the catheter into the portal vein. Clear mesenteric tissue was extracted and placed in DMEM supplemented with 2% antibiotic and antimycotic solutions. Sterile scissors were used to finely mince the tissue into ~1 mm3 pieces. The minced tissue was centrifuged at 2500 rpm for 10 min at RT and washed twice with PBS. For in vitro digestion, 0.25% collagenase IV was previously prepared and prewarmed at 37 C, tissue was resuspended in enzyme solution supplemented with 3mM CaCl2 at constant shaking for proper digestion. Following digestion for 30 min at 37 C, the tube was transferred to a biosafety cabinet, passed the digested tissue suspension through a 70 μm strainer placed into a sterile 50 ml tube. Equal amounts of sterile DMEM were passed through a strainer to inactivate the enzyme. The cell suspension was then centrifuged at 1200 rpm for 5 min at 4 °C and washed twice with PBS.

For sorting of LyECs, LyEC-specific primary antibody podoplanin (pdpn, 1:200) and CD31 (1:100) was added in the cell suspension with CD45 and 7AAD, and cells were incubated on ice for 30 mins. After washing, secondary antibody conjugated with fluorochrome was added at a 1:500 dilution and incubated on ice for 30 min. Thereafter, sorting was performed under sterile conditions using a BD FACS Aria, and the cells were collected in EGM-2 media. Collected cells were washed with PBS, resuspended in EGM-2 medium, and seeded on pre-coated fibronectin plates.

The cultured cells were incubated with nanoengineered VEGF-C particles for 30 min at 37 C in 5% CO2. After incubation, the cells were rinsed with PBS to remove the remaining nanoparticles. Cells were then fixed for 7 min at room temperature in the dark using a permeabilization and fixing kit (BD Cytofix/Cytoperm) and washed once with BD perm/wash buffer [7]. Staining was performed with DAPI (4,6-diamidino-2-phenylindole) at a 1:2000 dilution. Fluorescence of coumarin-6 labeled VEGF-C particle and DAPI-stained cells were imaged using an inverted fluorescence microscope (Evos microscope) in the green and blue channels, respectively. Fluorescence was quantified using the ImageJ software.

***Assessment of blood and lymphatic vessels by immunohistochemistry and immunofluorescence***

Tissues samples were fixed in 10% buffered formalin and processed. Sections of 7-μm-thick paraffin-embedded tissues were heat fixed and deparaffinized at 45 C and rehydrated in a descending ethanol series. Following antigen retrieval by heating for 8 minutes in a microwave with citrate buffer, sections were incubated for 20 minutes with peroxidase 1 solution to quench endogenous peroxidase. Protein blocking was done 3% BSA. Tissue slides were then incubated overnight at 4C in a humid chamber with anti-podoplanin mAb, and staining was completed using the HRP-conjugated mouse/rar/human detection kit and DAB chromogen as a substrate, according to the manufacturer’s instructions. Last, sections were counterstained with Hematoxylin and eosin for 1 minutes. The slides were mounted with a coverslip using Mounting Media. In immunofluorescence, after primary antibody incubation, secondary antibody attached to fluorochrome is added for 1 hr at RT and slides were mounted with Vectashield mounting media with DAPI. Six fields from each slide were randomly selected, and photographs were taken using an inverted fluorescent microscope (Nikon Instruments, Inc.) and quantified using ImageJ software. Details of the antibodies used are provided in S Table 4.

***Immune cell quantification using Flow Cytometry***

Cells were isolated from Mesenteric lymph node using enzymatic digestion by collagenase type IV at 37 C for 10 mins and single cell suspension was prepared using 40-micron sterile filter. Rest of the tissue was was passed through the filter using 5 ml syringe plunger. Single cell suspension was washed with PBS and counted. Cells from blood was isolated using RBC lysis buffer. 9 ml of RBC lysis buffer 1X was added to 1 ml of blood and centrifuged at 1500 rpm, 25 C for 5 min. cells were washed with PBS and counted. Half million cells were incubated with antibodies specific for T cell subsets and dendritic cells and incubated for 30 min to 1 hr in dark at 4C. 1 lakh events were acquired for each experiment.

***Detection of TNF-α and endotoxins in the Ascitic fluid and Systemic Circulation***

TNF-α levels were assessed using a TNF-α ELISA kit (Thermo Fisher Scientific, Massachusetts, USA) as recommended by the manufacturer’s protocol. Endotoxin levels in the serum were assessed using a chromogenic kinetic limulus ameobocyte lysate assay kit, following the manufacturer's instructions **(**Thermo Fisher Scientific, Massachusetts, USA).

***RNA extraction and RT-PCR***

RNA extraction and RT-PCR were performed on excised mesenteric tissues stored in an RNA buffer. Total RNA was isolated using a Nucleopore kit, according to the manufacturer’s instructions. RNA was quantified at 260/280 nm using a Nanodrop 2000 spectrophotometer (Thermo Scientific). First-strand cDNA was synthesized from 1µg of total RNA using reverse transcriptase (Thermo Fisher Scientific Verso cDNA synthesis kit) according to the manufacturer’s instructions. Quantitative real-time PCR was performed using SYBR green PCR master mix (Fermentas Life Sciences) on a ViiA7 instrument PCR system (Applied Biosystems, USA). The following cycling parameters were used: start at 95 °C for 5 min, denaturation at 95 °C for 30 s, annealing at 60 °C for 30 s, elongation at 72 °C for 30 s, and a final 5 min extra extension at the end of the reaction to ensure that all amplicons were completely extended and repeated for 40 amplification cycles. Relative quantification of the expression of relevant genes was performed using the ΔΔCt method after normalization to the expression of the housekeeping gene GAPDH. Primer sequences used are listed in S Table S5.

***Western Blotting***

Mesenteric tissues were crushed in liquid nitrogen, and 100 mg of tissue powder was added to 200 µL of RIPA lysis buffer (Merck, sigma 20-188). Homogenization was performed on ice until a clear solution was obtained. After centrifugation at 12,000 rpm for 20 min, the supernatant was collected in fresh microcentrifuge tubes and incubated on ice for 30 min. The protein content of the tissue lysate was measured using a BCA kit (Thermo Fisher Scientific, Waltham, MA, USA). Protein samples were denatured at 95°C for 5’ in Laemmli buffer. 60 µg of protein was loaded into each well and separated by 10% SDS-PAGE. The gel was run at 80 V for approximately 2 hr. Proteins were electroblotted onto activated PVDF membrane at 60 V for 2 hr. at 4°C, and the membrane was blocked in 5% BSA in Tris-buffered saline containing 0.05% Tween for 2 hr. Membranes were blotted with various primary antibodies i.e., VEGF-C and GAPDH overnight at 4°C, followed by the appropriate HRP-conjugated anti-rabbit and anti-mouse secondary antibodies for 2 hr. The membrane was then treated with the chemiluminescence ECL, and visualized on gel doc (Invitrogen, iBrightCL1500). Densitometry was performed using NIH software (ImageJ). Details of the antibodies used are provided in S Table 4.

**Supplementary Figures legend**

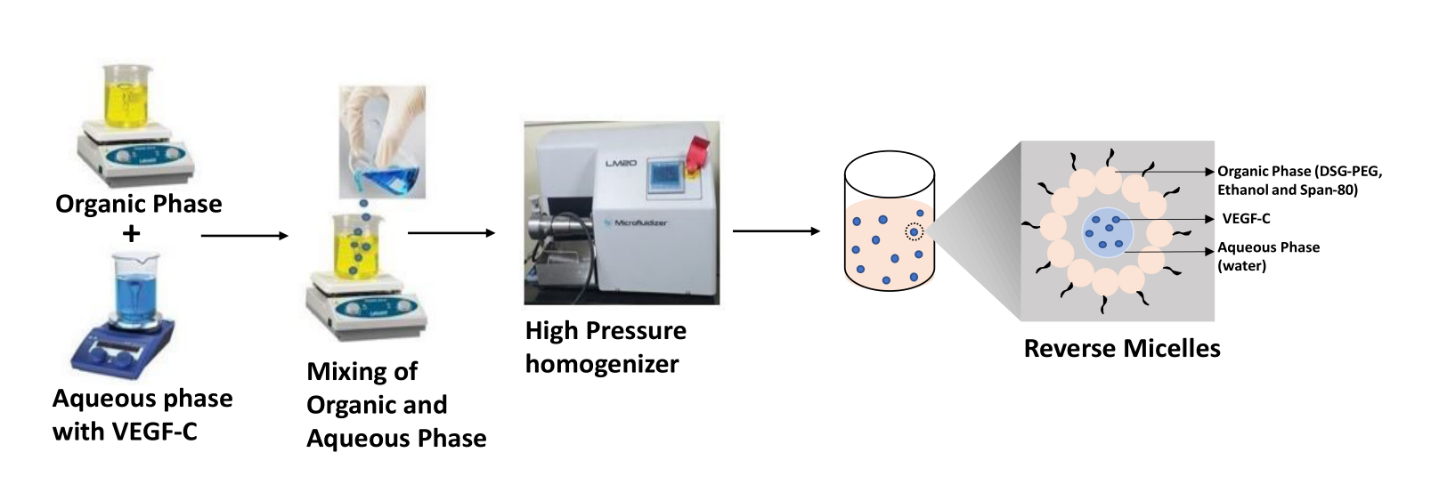

**Supp Figure 1:** **Schematic diagram for the preparation of reverse micelles:** Preparation of nanoengineered Distearoyl-rac-glycerol-PEG2K (DSG-PEG) based reverse micelles loaded with human recombinant VEGF-C (Cys56Ser) using homogenization.

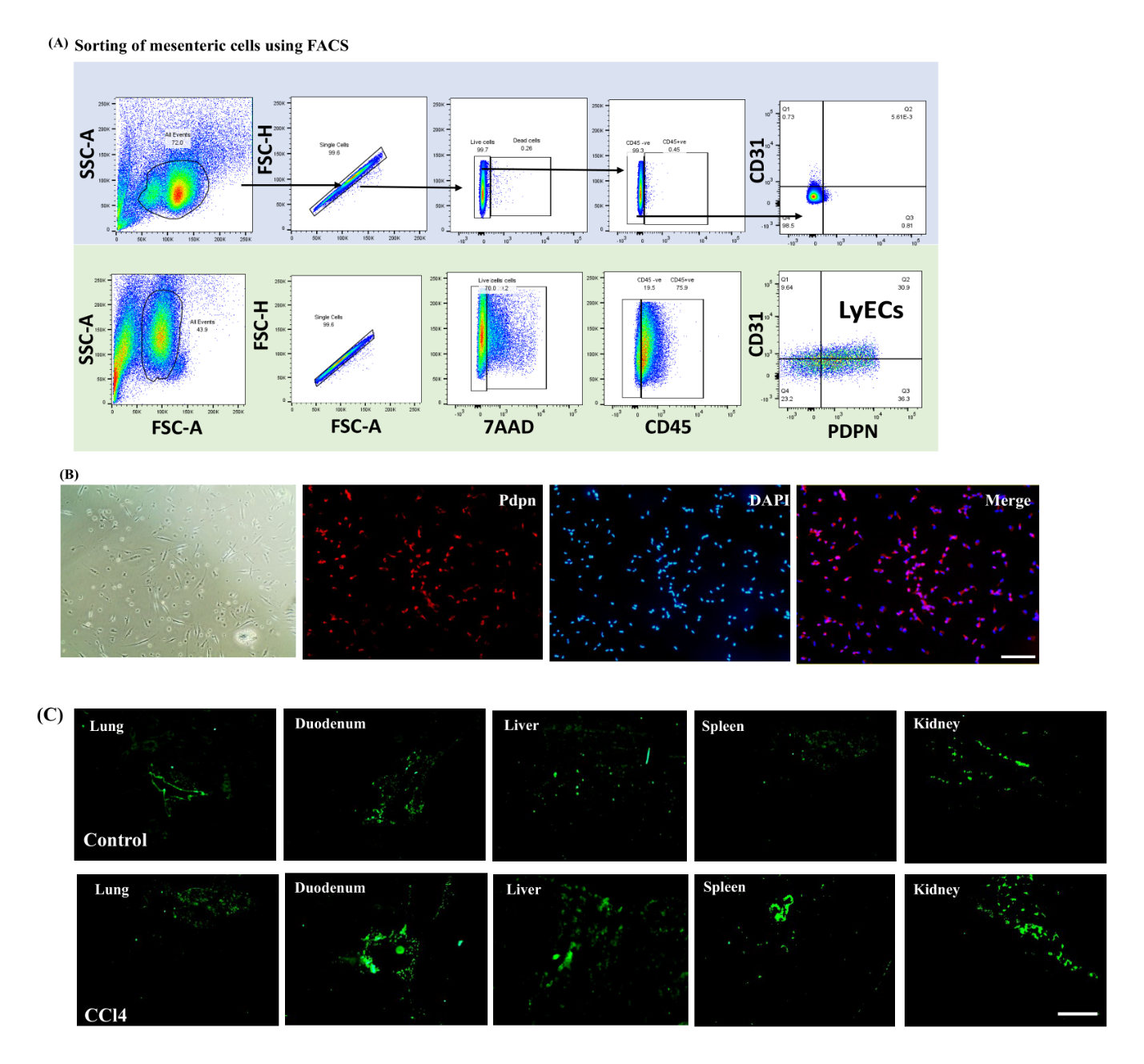

**Supp Figure 2:** **Sorting and culture of mesenteric lymphatic endothelial cells (LyECs). (A)** Primary cells were isolated from rat mesentery and labelled with CD31 and Pdpn antibodies for sorting LyEC using FACS. Dot plot graph showing the gating for sorting LyECs positive for CD31 and Pdpn. (**B)** Culture of sorted LyECs on the fibronectin-coated dish. Magnification 20x. (**C)** Ex vivo bio-distribution of coumarin-6 tagged E-VEGF-C in control and CCl4 rats after 2 hours of E-VEGF-C administration via the oral route. Scale bar: 500µm.

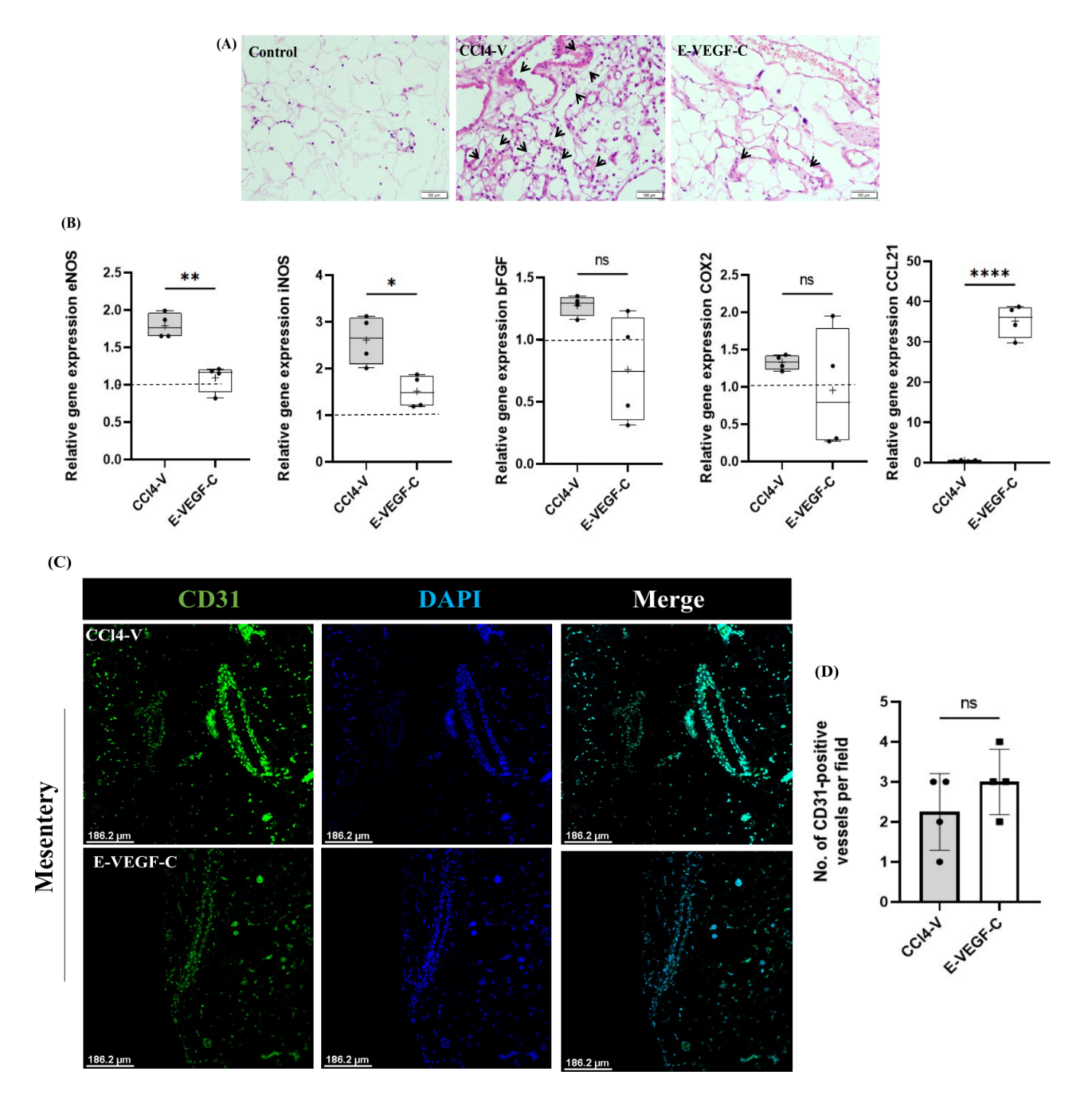

**Supp Figure 3: (A)** Hematoxylin and Eosin staining of control, CCl4-V, and E-VEGF-C mesentery sections. E-VEGF-C treated rats showed less immune cell infiltration and inflammation compared to the CCL4-V rat mesentery section indicated by the arrow. **(B)** Relative gene expression of eNOS, iNOS, bFGF, COX2, and CCL21 in mesentery tissue with mesenteric lymph node extract in all study groups. The dotted line represents control. n=4 each. Data is expressed as mean ± SD. **(C)** Immunofluorescence staining of an antibody recognizing CD31 in mesentery tissue section. DAPI is used for nuclear staining. (**D)** Quantitative analysis of the CD31+ blood vessels in mesentery of CCl4-V and E-VEGF-C treated. n=4 each. Data is expressed as mean ± SD. ns= non-significant, ‘*’ represents p< 0.05 and ‘**’ represents p< 0.01, and ‘***’ represents p< 0.001

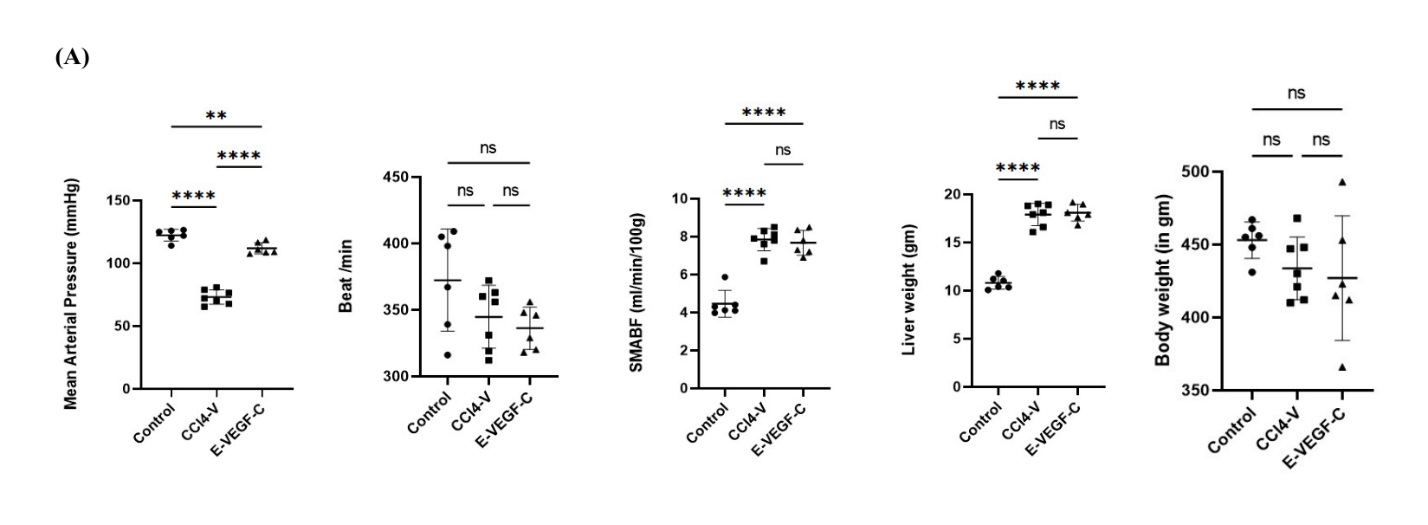

**Supp Figure 4:** Bar Diagrams showing hepatic hemodynamic and physiological parameters such as Mean Arterial Pressure (MAP), Beats/min, Superior Mesenteric Artery Blood Flow (SMABF), Liver weight, and Body weight in all study groups. n=6 each. Data is expressed as mean ± standard deviation. One Way ANOVA with post hoc Tukey’s test was performed ‘*’ represents p< 0.05 and ‘**’ represents p< 0.01, and ‘***’ represents p< 0.001

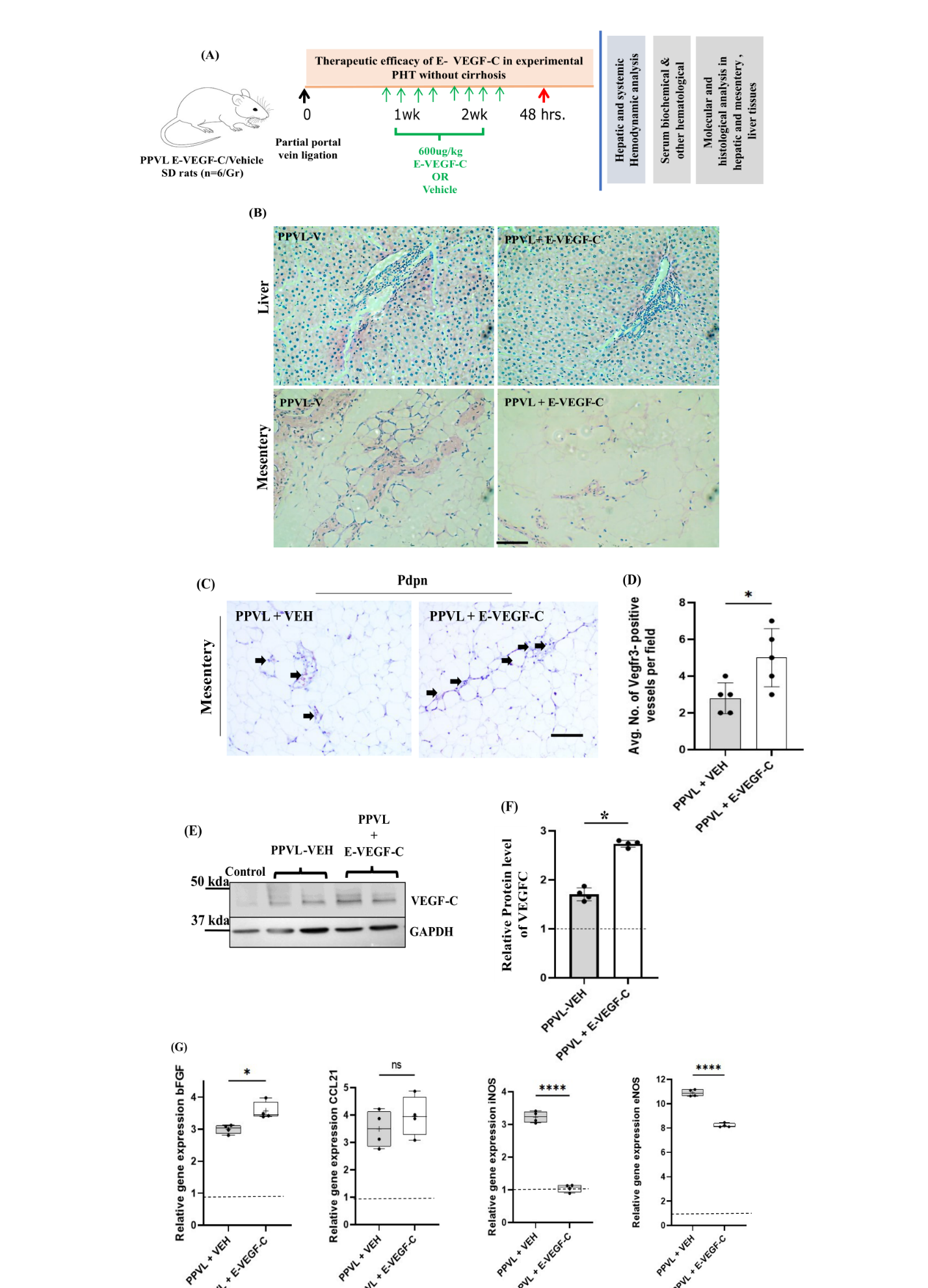

**Supp Figure 5:** **Effect of E-VEGF-C treatment on mesenteric lymphatic vessels in non-cirrhotic rat model of portal hypertension.** (**A)** Schema of the *in vivo* studies. Partial Portal Vein Ligation (PPVL) was done in rats to develop portal hypertension without cirrhosis. One week after surgery, rats were treated with E-VEGF-C 600ug/kg on alternate days for 2 weeks. n=6 each group. (**B)** Hematoxylin and Eosin staining of liver and mesentery tissue section of PPVL rats treated with Vehicle or E-VEGF-C. Scale bar: 500um. (**C)** Immunohistochemistry staining for antibody recognizing VEGFR3 in the mesenteric tissue sections of PPVL-Veh and PPVL+ E-VEGF-C. Scale bars represent 500μm. Arrow indicated at VEGFR3+LVs. (**D)** Quantitative analysis of the same section. VEGFR3+ LVs were counted and plotted. n=5 each. Data is expressed as mean ± SD. Unpaired two-tailed t-test were performed. *P*=0.0352. (**E)** The expression of human VEGF-C protein was measured using Western Blotting in control, PPVL-Veh, and PPVL+E-VEGF-C treated rats. **(F)** Quantitative analysis of the western blot is represented in bar graph. Dotted lines represent control. n=4 each. Data is expressed as mean ± SD. (**G)** Relative gene expression of bFGF, CCL21, iNOS, and eNOS and in mesentery tissue with mesenteric lymph node extract of PPVL-Vehicle and PPVL-E-VEGF-C rats. Dotted lines represent control. n=4 each. Data is expressed as mean ± SD. One Way ANOVA with post hoc Tukey’s test was performed. ‘*’ represents p< 0.05, ‘**’ represent p<0.01 and ‘***’ represents p< 0.001.

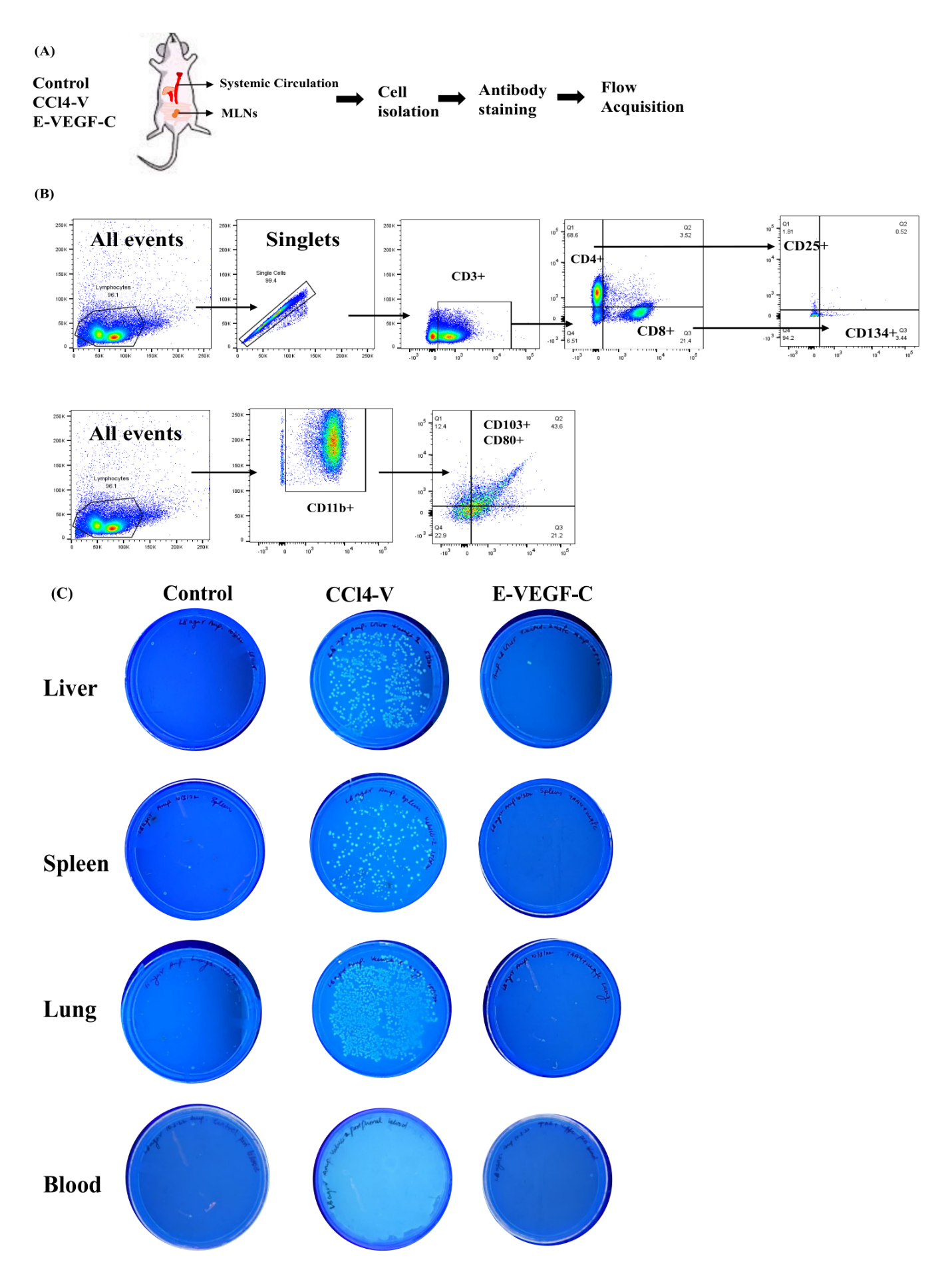

**Supp Figure 6: (A)** Schema representation of the workflow. Cells were isolated from the mesenteric lymph node of control, CCl4-V, and E-VEGF-C treated rats and labelled with antibodies for T cell subsets and Dendritic cells. **(B)** Gating strategy for the flow cytometry quantification of T cell subsets and Dendritic cells. 1 lakh events were acquired for each experiment. **(C)** GFP labelled Salmonella typhimurium 10^9^ bacteria were orally given in control, CCl4 and E-VEGF-C treated rats and tissue were collected after 48 hrs. 100 mg of Liver, Spleen, and Lung were homogenized and plated on LB agar plate. Blood was collected and transferred to a blood culture bottle and incubated for 1 hr. in 37 C. 100 ul of blood was collected from the blood culture bottle using a sterile syringe and plated on LB agar plate. Plates were observed in UV transilluminator after 24 hrs. of incubation at 37 C.

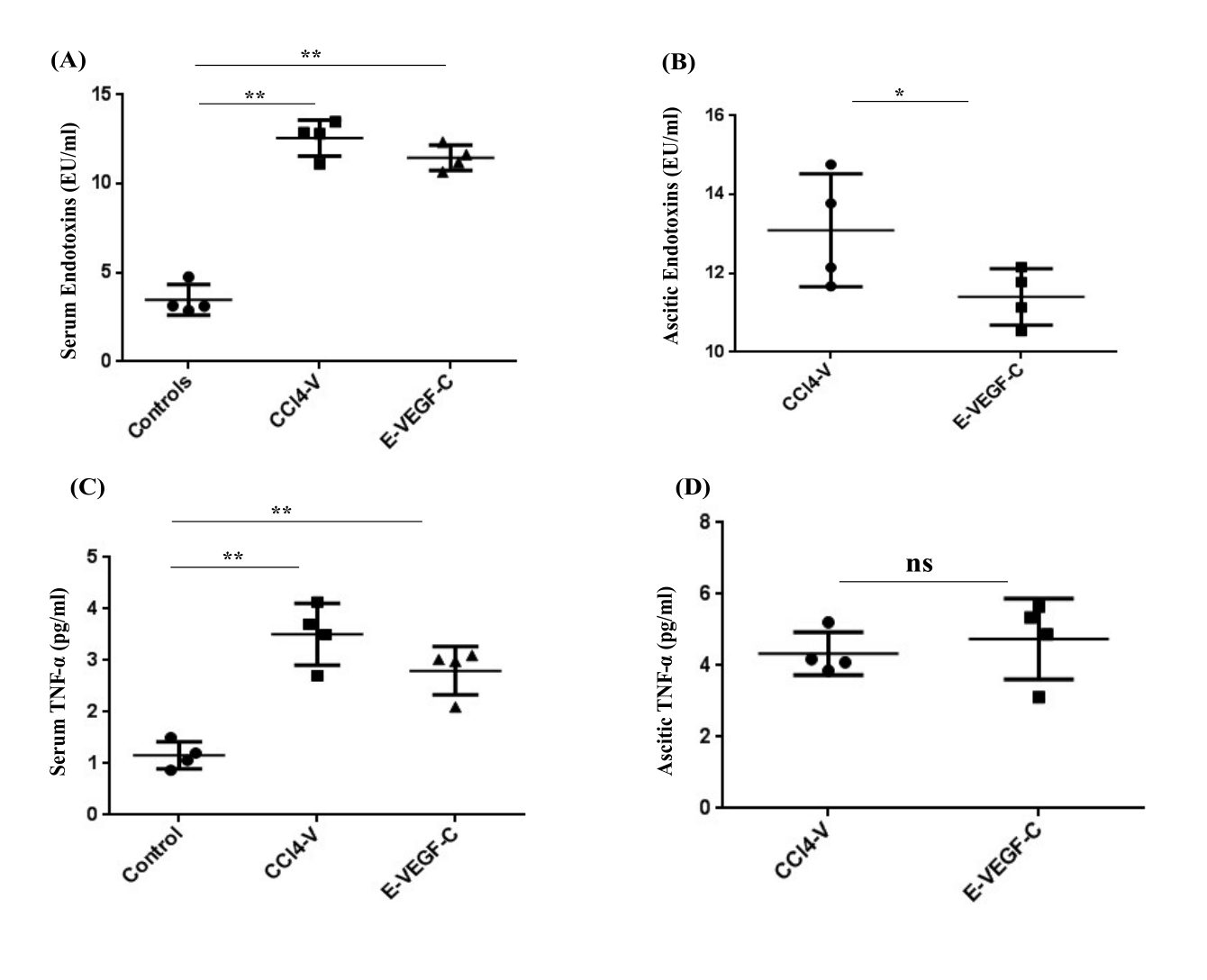

**Supp Figure 7:** Quantification of endotoxins and TNF-α in serum and ascitic fluid of control, CCl4-V, and E-VEGF-C rats. Dot plots showing levels of Endotoxins (EU/ml) **(A)** in serum and (**B)** ascitic fluid. Dot plots showing levels of TNF-α (pg/ml) **(C)** in serum and **(D)** in ascitic fluid. n=4 each. Data is expressed as mean ± SD. ‘*’ represents p< 0.05, and ‘**’ represents p<0.01.

**Supplementary Tables**

| **Properties** | **Time points (week)** | | | | |
| --- | --- | --- | --- | --- | --- |
|  | **0** | **1** | **2** | **3** | **4** |
| **Particle size (nm)** | **134.8 ± 0.47** | **135.4 ± 1.70** | **137.7 ± 0.98** | **141.4 ± 2.51** | **152.7 ± 0.30** |
| **Polydispersity index** | **0.126 ± 0.01** | **0.119 ± 0.01** | **0.104 ± 0.005** | **0.083 ± 0.017** | **0.078 ± 0.011** |
| **Zeta potential (mV)** | **-21.9 ± 1.24** | **-24.5 ± 0.79** | **-18.9 ± 1.70** | **-16.4 ± 0.153** | **-15.8 ± 0.47** |

**Supp Table 1.** Reverse micelles properties monitored during stability study

**Supp Table 2** R^2^ value obtained from different kinetic models

| **Kinetic models** | **R^2^ value** |
| --- | --- |
| Zero order | 0.999 |
| First order | 0.884 |
| Korsmeyer-Peppas | 0.973 |
| Weibull | 0.997 |
| Higuchi | 0.947 |
| Hixson-Crowell | 0.176 |

**Supp Table 3: Pathological Parameters of the cirrhotic animal study groups**

| **Parameters** | **Control** | **CCl4-V** | **E-VEGF-C** |
| --- | --- | --- | --- |
| **ALT (IU/L)** | 42.8 ± 2.3 | 64 ± 4.5 | 72 ± 5.6 |
| **Albumin (g/dL)** | 3.9 ± 0.45 | 1.2 ± 0.35 | 1.3 ± 0.41 |
| **Sodium (mmol/L)** | 145 ± 1.4 | 158 ± 2.1 | 148 ± 2.9 |
| **Urea (mg/dL)** | 51.5 ± 1.8 | 68.5 ± 1.9 | 49.4 ± 2.3 |
| **Creatinine (mg/dL)** | 0.76 ± 0.03 | 0.66 ± 0.08 | 0.85 ± 0.02 |
| **Fibrosis** | - | ≥5 | 5 |
| **Ascites (% of total animals)**   - **Mild** - **Moderate** - **Severe** |  | 0  50  50 | 50  50  0 |

**Supp Table 4: Antibody Panel**

| **Antibody** | **Brand** | **Catalogue/Clone** | **Fluorochrome** | **Application** | **Dilution** |
| --- | --- | --- | --- | --- | --- |
| Podoplanin | eBioscience | **eBio8.1.1**  **14-5381-82** | Unconjugated | Used for IHC, Whole mount and FACS | 1:200 for IHC  1:100 for W.M.  0.5ug/test for FACS |
| Goat Anti Rabbit IgG PE | Santa Cruz | sc-3739 | PE | Used for IF and IHC with vegfr3 | 1:500 |
| CD31 | RnD system | AF3628 | Unconjugated | Used for IF | 1:400 |
| CD45 | BioLegend | 202225 | Pacific blue | Used for FACS | 1:200 |
| 7AAD | BioLegend | 420403 | Dye | Used for FACS | 1:500 |
| Goat Anti mouse FITC | Merck-Millipore | AP308F | FITC | Used for IF | 1:500 |
| VEGFR3  (Host: Rabbit) | Thermofisher Scientific | PA1-37712 | Unconjugated | Used for IF | 1:100 |
| VEGF-C (Host: Mouse) | Invitrogen | MA5-26494 | Unconjugated | Used for western blotting | 1:500 |
| CD3 | BioLegend | 201413 | APC | Used in flow cytometry | 1:200 |
| CD4 | BioLegend | 201519 | PE-Cy7 | Used in flow cytometry | 1:100 |
| CD8 | BioLegend | 201715 | PerCP-Cy5.5 | Used in flow cytometry | 1:100 |
| CD25 | BioLegend | 202103 | PE | Used in flow cytometry | 1:100 |
| CD134 | BioLegend | 204508 | FITC | Used in flow cytometry | 1:100 |
| CD11b | BioLegend | 201817 | PE-Cy7 | Used in flow cytometry | 1:100 |
| CD103 | BioLegend | 205509 | APC | Used in flow cytometry | 1:200 |
| CD80 | BioLegend | 200205 | PE | Used in flow cytometry | 1:100 |
| GAPDH (Host: mouse) | Santa Cruz Biotechnology | SC-32233 | Unconjugated | Used for western blotting | 1:5000 |
| Anti-Mouse IgG Antibody | Santa Cruz Biotechnology | SC-5616102 | HRP conjugated | Used for western blotting | 1:10,000 |
| Anti-Rabbit IgG Antibody | Sana Cruz Biotechnology | SC-2357 | HRP conjugated | Used for western blotting | 1:10,000 |

IHC: Immunohistochemistry, IF: Immunofluorescence

**Supp Table 5: List of rat genes and primers used for qRT-PCR**

| **Gene** | **Forward Primer** | **Reverse Primer** |
| --- | --- | --- |
| **LyVE1** | GAAATGCAGACCCACAGAT | AACCCATCCATAGCTGCAAG |
| **Prox1** | GCATAAACCCCCAGACCCAA | AGCCAAGCTCACATCTCACC |
| **VEGF-C** | TGTATAGATGTGGGGAAGGA | ACAACGTCTTGCTGAGGTAA |
| **VACM-1** | AGTGTGAATCGAAAACCGAA | GATGCAAAGTAGAGTGCAAG |
| **VE-Cad** | ACAACTTCCCCATCTTTACTC | ATATTTGGTCATCCGGTTCT |
| **MHCII** | GAAAGGGGACCTGGACTT | TGGCACACAGAGTATGG |
| **CD86** | TTGGGAATCCTTTTCTCGGTG | TTTGAGCCTTTGTGAACGGG |
| **bFGF** | CCAGTTGGTATGTGGCACTG | CAGGGAAGGGTTTGACAAGA |
| **iNOS** | ACC TAC TTC CTG GAC ATC AC | ACC CAA ACA CCA AGG TCA TG |
| **eNOS** | TGACCCTCACCGATACAACA | CGGGTGTCTAGATCCATGC |
| **COX2** | CTG TAT CCC GCC CTG CTG GTG | ACT TGC GTT GAT GGT GGC TGT CTT |
| **CCL21** | GGGACTGAACAGACAGACTCCAAG | GGTTGAAGCAGACAAGGGTGTG |
